# An ensemble prediction model for COVID-19 mortality risk

**DOI:** 10.1101/2022.01.10.22268985

**Authors:** Jie Li, Xin Li, John Hutchinson, Mohammad Asad, Yadong Wang, Edwin Wang

## Abstract

**Background:** It’s critical to identify COVID-19 patients with a higher death risk at early stage to give them better hospitalization or intensive care. However, thus far, none of the machine learning models has been shown to be successful in an independent cohort. We aim to develop a machine learning model which could accurately predict death risk of COVID-19 patients at an early stage in other independent cohorts.

**Methods:** We used a cohort containing 4711 patients whose clinical features associated with patient physiological conditions or lab test data associated with inflammation, hepatorenal function, cardiovascular function and so on to identify key features. To do so, we first developed a novel data preprocessing approach to clean up clinical features and then developed an ensemble machine learning method to identify key features.

**Results:** Finally, we identified 14 key clinical features whose combination reached a good predictive performance of AUC 0.907. Most importantly, we successfully validated these key features in a large independent cohort containing 15,790 patients.

**Conclusions:** Our study shows that 14 key features are robust and useful in predicting the risk of death in patients confirmed SARS-CoV-2 infection at an early stage, and potentially useful in clinical settings to help in making clinical decisions.

## Background

The global COVID-19 pandemic is putting high pressure on healthcare systems around the world [1-3]. Most of people infected with the SARS-CoV-2 have mild disease and self-limiting, but there still a significant proportion of patients developed severe disease or even death [4, 5]. In epidemic areas, the shortage of medical resources may lead to an increase in mortality [6]. Therefore, it is important to distinguish patients at high risk of severe illness or mortality from others in the early stages of disease development.

There have been several researchers contributing to the areas of mortality risk prediction for patients. In May 2020, Yan et al. selected three biomarkers that predict the mortality of individual patients more than ten days in advance through machine learning tools [7]. However, Yan et al. gathered samples from a cohort of only 485 patients with confirmed SARS-CoV-2 infection may not be sufficient, and their mortality predictive model may not perform well in other cohorts [8]. Comorbidity, biomarkers for kidney disease and other clinical characteristics have also been associated with the severity of a patient’s disease, according to previous studies [9-12]. Liang et al. proposed a deep learning-based survival model can predict the risk of COVID-19 patients developing critical illness based on clinical characteristics at admission [13]. The deficiency of this study is that the features selected may not sufficient to reflect the patient’s condition, which might be the reason for the differences in the performance of different validation sets [13]. Based on clinical information, Altschul et al. proposed a novel severity score to assess the severity of patients infected with the SARS-CoV-2 [14]. Patients were classified into low (0-3), moderate (4-6), and high (7-10) COVID-19 severity scores. A ROC curve analysis showed that the AUC of the derivation cohort was 0.824 and the AUC of the validation cohort was 0.798 [14].

Except these examples, thus far, lots of similar efforts have been made by others. Recently, Wynants et al. conducted a comprehensive and systematic review of 145 prediction models from 107 studies, with a brief summary of the features (predictors) used by these models [15]. The key message from this analysis was that none of the models can be validated independently (i.e., their predictions failed when validating in an independent cohort), in another word, none of the predictive models developed in the COVID-19 domain could be used in clinics for decision marking. The prediction power of most of the models was similar to that of flipping a coin [15]. In this study, we look forward to developing a machine learning model which could accurately predict mortality risk of COVID-19 patients at an early stage in other independent cohorts.

## Methods and Materials

The flow chart of our prediction method is shown in Figure 1. We first group the features and preprocess them, including processing extreme values and imputing missing values. Then, feature selection is carried out for constructing ensemble model (EM). Five base models: Gradient Boosted Decision Tree (GBDT), Extreme Gradient Boosting (XGBoost) [16], Random Forest (RF), Logistic Regression (LR), and Support Vector Machine (SVM) are used to select features. Features selected by more than half (3 or more) of the base models are used to construct the the EM. Finally, performance of the EM and base models is compared and validated on independent datasets.

**Figure 1.**
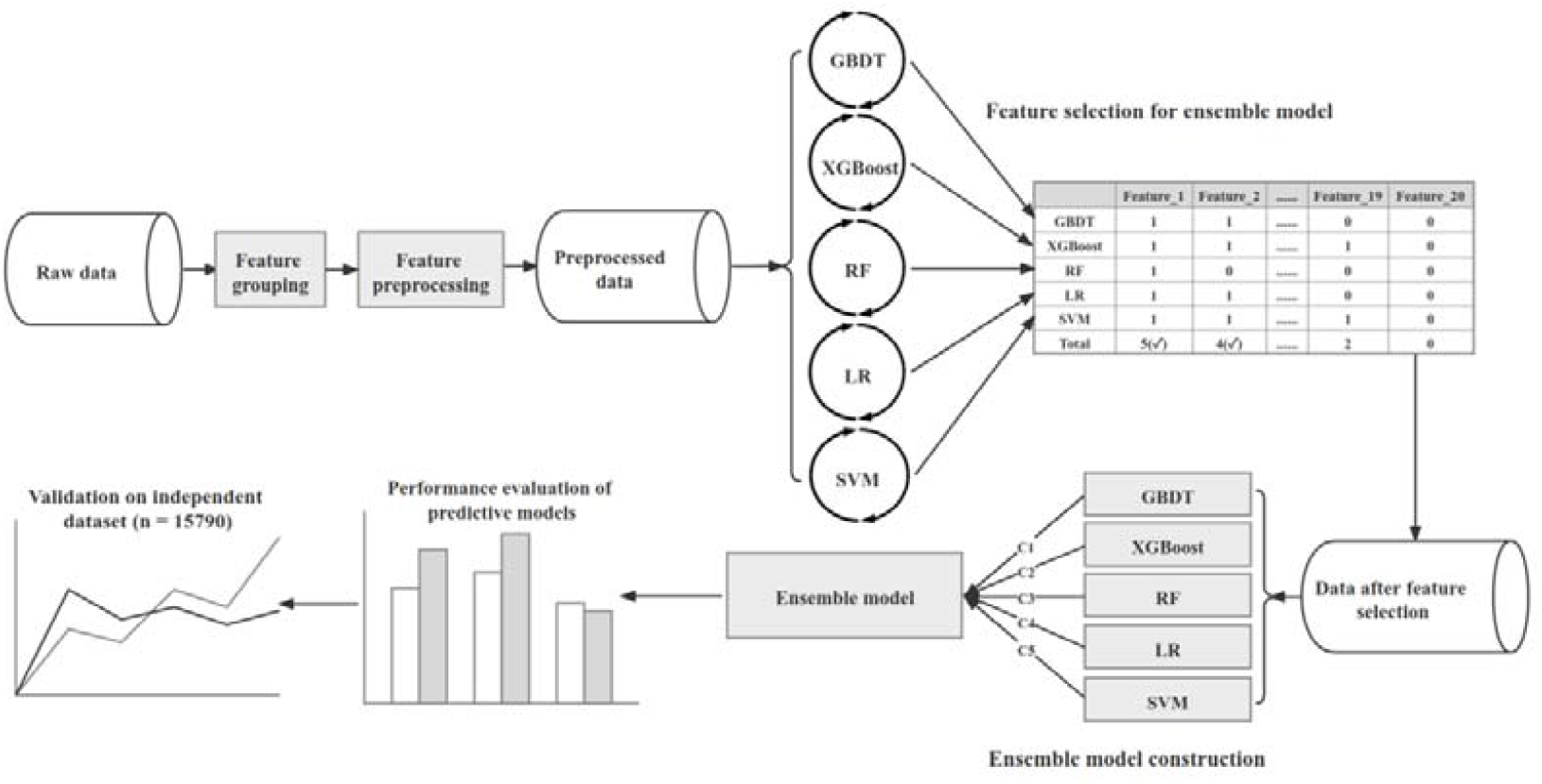
The flow chart of our prediction method

### Data sets

Two datasets are used in the study: Cohort 1 and Cohort 2. Cohort 1 with 4,711 COVID-19 patients (1,148 deaths) is from a recent study [14], which was collected from March 1, 2020, to April 16, 2020. All patients in cohort 1 are hospitalized, and their clinical features are obtained at admission[14]. Clinical features include patient’s age, mean arterial pressure, oxygen saturation, etc., and the details and statistical information of these clinical features are shown in Table 1 and Supplementary Table 1. According to the types and meanings of clinical features (Table 1), numerical variables (features) that can directly reflect the physiological conditions of patients are selected features of models. These clinical features include age, oxygen saturation (OsSats), temperature (Temp), mean arterial pressure (MAP), D-dimer (Ddimer), platelets (Plts), international normalized ratio (INR), blood urea nitrogen (BUN), creatinine, sodium, glucose, aspartate aminotransferase (AST), alanine aminotransferase (ALT), white blood cells (WBC), lymphocytes (Lympho), interleukin-6 (IL-6), ferritin, C-reactive protein (CrctProtein), procalcitonin and troponin. If COVID-19 patient dies, his or her label is set to 1, otherwise it will be set to 0. We select features and trained our models on Cohort 1. Cohort 2 is an independent validation data containing 15,790 COVID-19 patients from UK biobank [17, 18]. The statistical results of clinical features and population structure of this data are shown in Supplementary Table 2. The mortality rate is 4.21% (664/15790) in Cohort 2. We selected hundreds of features (which are identical to or functionally related to the features selected in Cohort 1) from Cohort 2, and divided them into 55 functionally related features (Supplementary Table 3). These features included age, blood pressure-related features (such as hypertension), kidney function related features (such as creatinine), inflammation related features (such as monocyte), and so on.

**Table 1.**
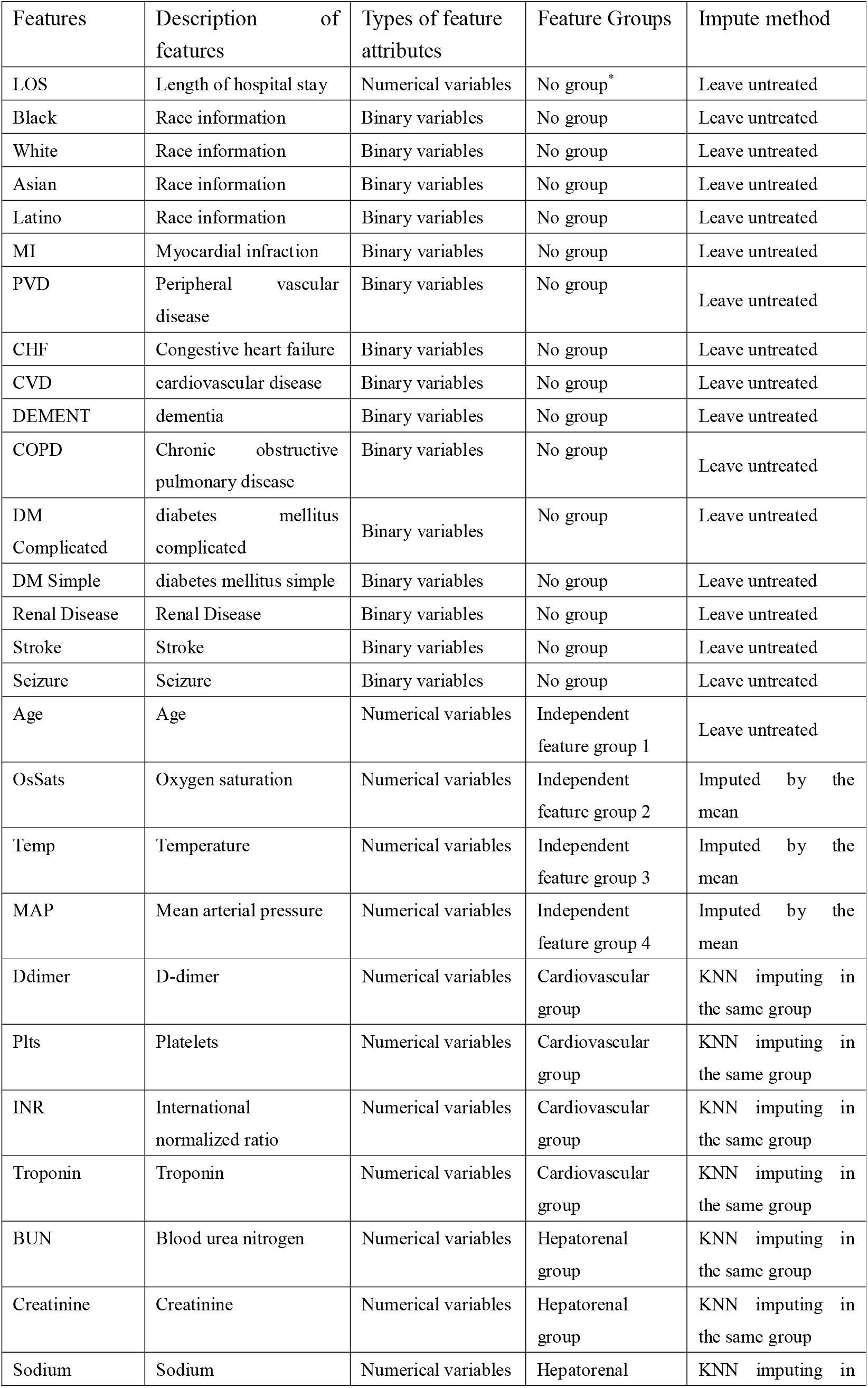
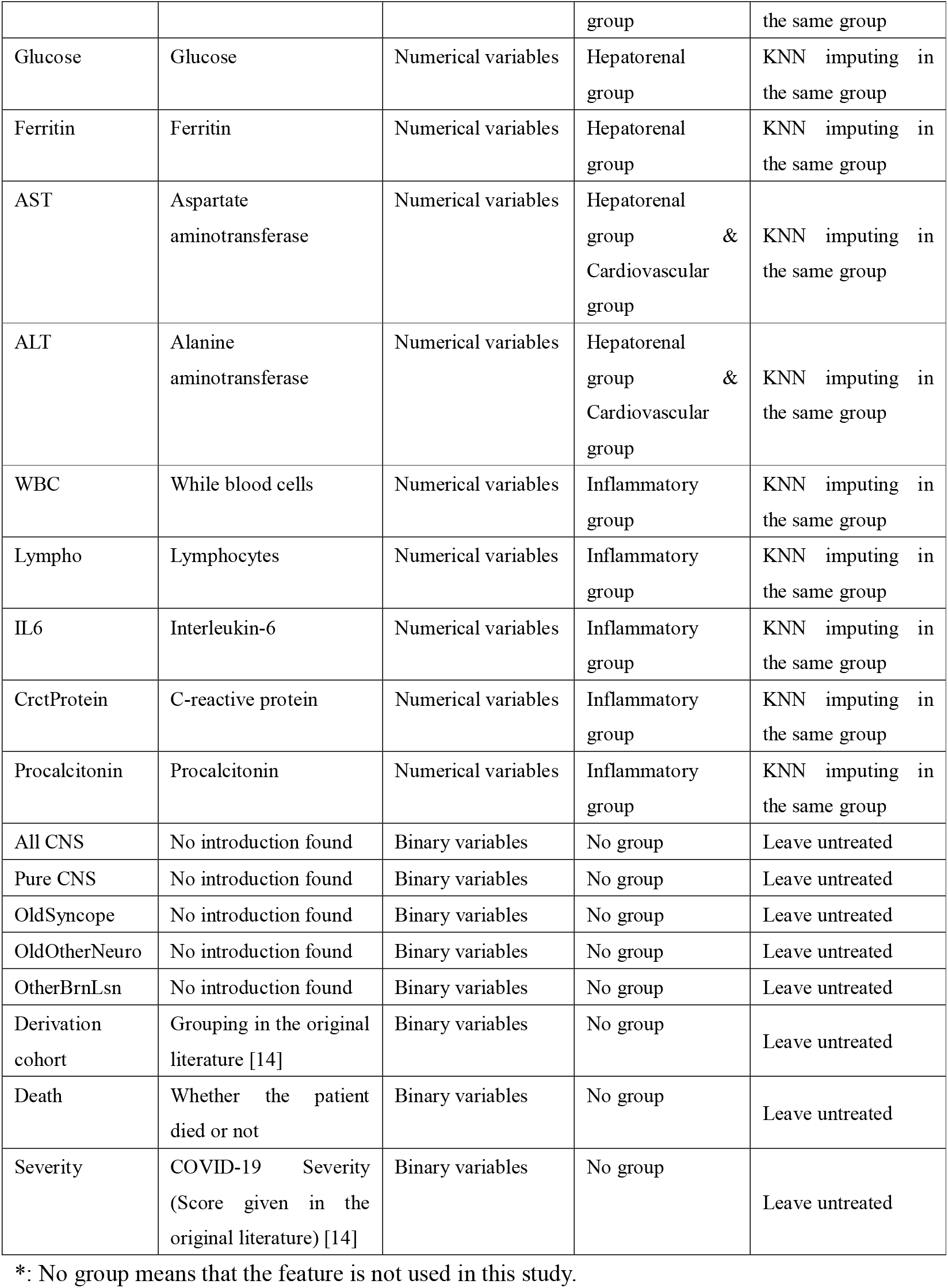
Clinical features of patients infected with SARS-CoV-2 in the cohort 1

### Feature grouping and feature preprocessing

The presence of missing/error data will reduce the performance of the predictive model. Therefore, we developed a novel feature grouping and preprocessing method to deal with missing/error data. We hypothesized that features closely related to patient’s physiological conditions are better indicators associated with death risk. Thus, a key concept in this study is to select features and group them based on if they could indicate a particular aspect of the patient’s physiology, but not strictly based on medical definitions. By doing so, age, OsSats, Temp, and MAP were divided into independent feature groups. Independent features are those that are not significantly associated each other. Other clinical features such as BUN, creatinine, glucose, sodium, ferritin, aspartate aminotransferase (AST), alanine aminotransferase (ALT) could indicate the conditions of liver or kidney [19-23], therefore, they were divided into the hepatorenal group. By the same token, IL-6, CrctProtein, Lympho, WBC, and procalcitonin are all inflammation-related features [24, 25], we combined these features into the inflammatory group. Troponin, AST, ALT, Ddimer, Plts, and INR could be associated with cardiovascular conditions [26-29], which were combined into the cardiovascular group. The detail of feature grouping is shown in Table 1.

One of the challenges of using clinical data is that clinical data often have missing values and error values (missing values are represented by 0 in the original dataset). To overcome this shortage, we developed a novel imputation pipeline to preprocess these features. First of all, we dealt with the extremums of some features. For each feature, we calculated its 95th percentile as the cut-off value (called cutoff95) and replaced feature value using cutoff95 if the feature value of certain patient is greater than cutoff95. The maximum of these features in the original dataset and the selected cutoff95 are shown in Table 2.

**Table 2.** The maximum in the original data and the selected cutoff95

| Feature | Maximum | Cutoff95 |
| --- | --- | --- |
| Ddimer | 20.00001 | 20.00001 |
| Plts | 1226 | 433 |
| INR | 17.0001 | 1.7 |
| BUN | 301 | 97 |
| Creatinine | 31.66 | 7.35 |
| Sodium | 170.001 | 153 |
| Glucose | 1000.001 | 423.6 |
| AST | 10000 | 159 |
| ALT | 3228 | 116 |
| WBC | 219.7 | 16.9 |
| Lympho | 209.1 | 2.4 |
| IL6 | 111040 | 372.74 |
| Ferritin | 100000 | 4508.55 |
| CrctProtein | 100.0001 | 34.3 |
| Procalcitonin | 50.0001 | 12.52 |
| Troponin | 9.56 | 0.21 |

For the independent feature group, the missing values and obvious error values (temperature 50 °C and −17.78 °C) were imputed with the mean value of the feature. We take the temperature feature as an example to illustrate how to impute the missing temperature value. First, dataset was divided into two groups (death and survival groups) according to whether the patient died or not, the mean temperatures of the two groups of patients were calculated, respectively, after removing 0 and obvious error values (the mean temperature of death group was defined as M1, the mean temperature of survival group was defined as M2). Then, we imputed the missing and error values within the group using the mean of each group (M1 for the death group, M2 for the survival group).

For other feature groups (i.e., inflammatory group, etc.), the missing values were imputed using k-nearest neighbor method (KNN, k=3). We take WBC in inflammatory group as an example to illustrate how to impute the missing WBC value. First, patients were divided into two groups (death and survival groups) according to whether the patient died or not. Then, patients in the same group were clustered using KNN according to 4 features: Lympho, IL6, CrctProtein, Procalcitonin. If WBC value of certain patient is missing, it is imputed by average WBC value of three nearest patients to the patient. For AST and ALT, we imputed their missing values using hepatorenal group, since both AST and ALT are associated with both hepatorenal conditions and cardiovascular conditions, but they are more associated with hepatorenal features.

### Base model parameter settings

In order to select valuable features and develop an EM which could take advantages of several base models such as Gradient Boosted Decision Tree (GBDT), Extreme Gradient Boosting (XGBoost) [16], Random Forest (RF), Logistic Regression (LR), and Support Vector Machine (SVM), we first conducted experiments in Cohort 1 and set the best parameters for five base models, respectively. For RF, GBDT, and XGBoost, we adjusted the number of decision trees (n_estimators), and for XGBoost, we also adjusted the maximum depth (max_depth) and the subsample ratio of features (colsample_bytree) to control over-fitting or under-fitting when constructing each tree. For the SVM model, we chose the radial basis function (RBF) (kernel) as the kernel function, and the regularization parameter C (C) was set to 0.7 to reduce overfitting. Since SVM favors the majority class on unbalanced datasets, we adjusted the weights of the two classes inversely proportional to the frequency of the classes (class_weight) in the dataset. In addition, z-score was used to standardize the data before input into LR and SVM models due to the characteristics of the algorithm. The detail of parameters of five base models is shown in Table 3.

**Table 3.** Parameter settings of five base models

| GBDT | XGBoost | RF | LR | SVM |
| --- | --- | --- | --- | --- |
| random_state = 10<br>learning_rate = 0.1<br>n_estimators = 110 | random_state = 10<br>learning_rate = 0.1<br>n_estimators = 150<br>max_depth = 12<br>colsample_bytree=0.3<br>use_label_encoder=False | random_state = 10<br>n_estimators=200 | random_state=10 | random_state = 10<br>kernel='rbf'<br>class_weight='balanced'<br>C=0.7<br>probability=True |

### Feature selection for ensemble model

Redundant features could be detrimental to predictive models to make correct predictions. Therefore, we screened out most valuable features from the clinical features to improve the performance of our predictive models. The feature selection process is divided into two steps. In the first step, we select high performance feature set for 5 base models, respectively, from the 20 features in Table 1. In the second step, we combine feature sets of five base models to form the final selected feature set.

Genetic algorithm (GA) is used to select feature set [30], which is a heuristic search algorithm that simulates the process of natural selection. In the feature selection process, the area under the ROC curve (AUC) of each base model is taken as the objective function, each individual in GA represents a set of features, consisting of a binary string called a chromosome, and multiple individuals constitute a population. In each generation, a subset of individuals with the highest fitness (maximizing the objective function) goes into the next generation. In this way, we finally select a feature set that makes the predictive model get the best performance.

Taking the feature selection for GBDT as an example, the population size is set to 40, and each chromosome is encoded into a binary string of length 20. Each position of the chromosome represents whether the corresponding feature is selected or not. We used the elite-tournament method[31] as a selection operator to select the chromosomes with the highest fitness in the population. The single-point crossover operator is chose as the offspring chromosome recombination method, the crossover probability is set to 0.7 and the probability of offspring mutation is the reciprocal of chromosome length. According to the above settings, after running 200 generations, the high performance feature set is selected for GBDT.

We also select high performance feature set for other base models. Thus each base model has a high performance feature set. We combine these feature sets, and select features that appear in more than half (3 or more) of the feature sets to form the final selected feature set.

### Ensemble model construction

In order to construct an EM, which could take advantages of the prediction results of several models, we chose the above five models as base models to construct our EM. Similar to the feature selection method mentioned above, we also used GA to find a set of coefficients C as the weight of the prediction results of the five base models. The prediction results of EM (prob_em_) for patients are the weighted average of the prediction results of each base model (prob_i_), as defined below:

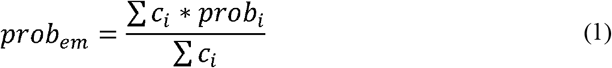

We used 0.5 as the threshold, and patient whose prediction result is higher than the threshold is predicted as dead. To obtain this set of coefficients C using the GA, fitness score is calculated using the AUC of the EM. We code each set of coefficients for five base models as a binary string (i.e., a chromosome), which length is 70. The number of individuals (chromosomes) in the population is set to 40. Other parameters of GA, such as selection operators and crossover operators, are the same as those used in feature selection process.

### Performance evaluation of predictive models

In this study, we evaluate the prediction performance of different models using accuracy, area under ROC curve (AUC), precision and recall. The definitions of accuracy, precision and recall are as followings:

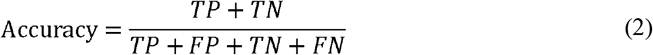

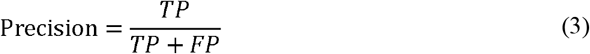

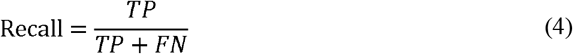

Here, TP, FP, TN, and FN represent the number of true positive, false positive, true negative and false negative, respectively. In this study, the death patient is positive sample. The predictive model gives the probability of death for each patient, and we set a threshold of 0.5, above which the patient is considered to be dead. Using clinical data composed of selected features as inputs and patient status (alive or death) as labels, we performed half-half cross-validations 100 times for each model (including EM and base models) on the entire cohort (Cohort 1, n=4,711). Specifically, we first randomly chose half of the samples (2,355 patients) as training set and the other half (2,356 patients) as test set from Cohort 1. Then we trained EM and base models on training set and tested the performance of these models on test set. Subsequently, training set and test set was exchanged (that is, the former training set was the test set and the former test set was the training set), and these models were retrained and tested. At the same time, prediction results of these models were saved. Above process repeats 100 times, average values and standard deviations (SD) of accuracy, AUC, precision and recall of EM and five base models were calculated.

### Validation on independent dataset

Cohort 2 is an independent cohort containing 15,790 patients (664 patients died and 15,126 patients survived). Since Cohort 2 did not fully contain the features in Cohort 1, we selected 55 features in Cohort 2 that were identical or functionally related to features selected from Cohort 1 (as shown in Supplementary Table 3 in this study) for further analysis. The missing data in Cohort 2 is imputed according to the method used in Cohort 1. In order to validate the performance of different models in Cohort 2, we first used GA to select a feature set that was most related to the mortality risk of patients from 55 features. Cohort 2 and Cohort 1 are different in several aspects, such as mortality rate, age distribution and so on (Supplementary Figure 1, Supplementary Tables 1 and 2). For comparison purposes, next, we selected a subset of patients aged 50 to 84 years from Cohort 1 (Subset 1), since the patients in Cohort 2 ranged in age from 50-84 years. As with Cohort 1, the data in subset 1 was also half-and-half cross-validation for 100 times. Subsequently, we randomly sampled a subset of patients (subset 2) with a similar percentage of patients surviving in subset 1 from Cohort 2 and run a half-and-half cross-validation on subset 2. The process also repeats 100 times. Finally, we test different models on subset 1 and subset 2 and calculated their accuracy, AUC, precision and recall.

## Results

We first selected feature set for each base model in Cohort 1 according our method, respectively. Feature sets of different base model were shown in Supplementary Table 4. Then, 14 key features were chose as the final feature set, which are Age, OsSats, MAP, Ddimer, Glucose, WBC, Lympho, IL6, CrctProtein, Procalcitonin, Troponin, Plts, INR and ALT. Finally, EM was constructed and the coefficients of base models of the EM calculated by GA were 0.39620338 (GBDT), 0.9574559 (XGBoost), 0.26222304 (RF), 0.0315571 (LR), and 0.24549838 (SVM), respectively. Mean values and standard deviations (SD) of accuracy, AUC, precision and recall of EM and five base models were shown in Table 4. Experimental results indicated that feature preprocessing and selection significantly improved the performance of the predictive models. In addition, the EM reached the best performance in unprocessed and preprocessed data, which showed the robustness of the EM. We also conducted an experiment in Cohort 1 using different imputation methods and test the performance of different models. Experimental results on Cohort 1 suggested that KNN imputation method was best and improved the performance of the predictive models than simply replacing missing data with 0 in the original data (as shown in Supplementary Table 5).

**Table 4.**
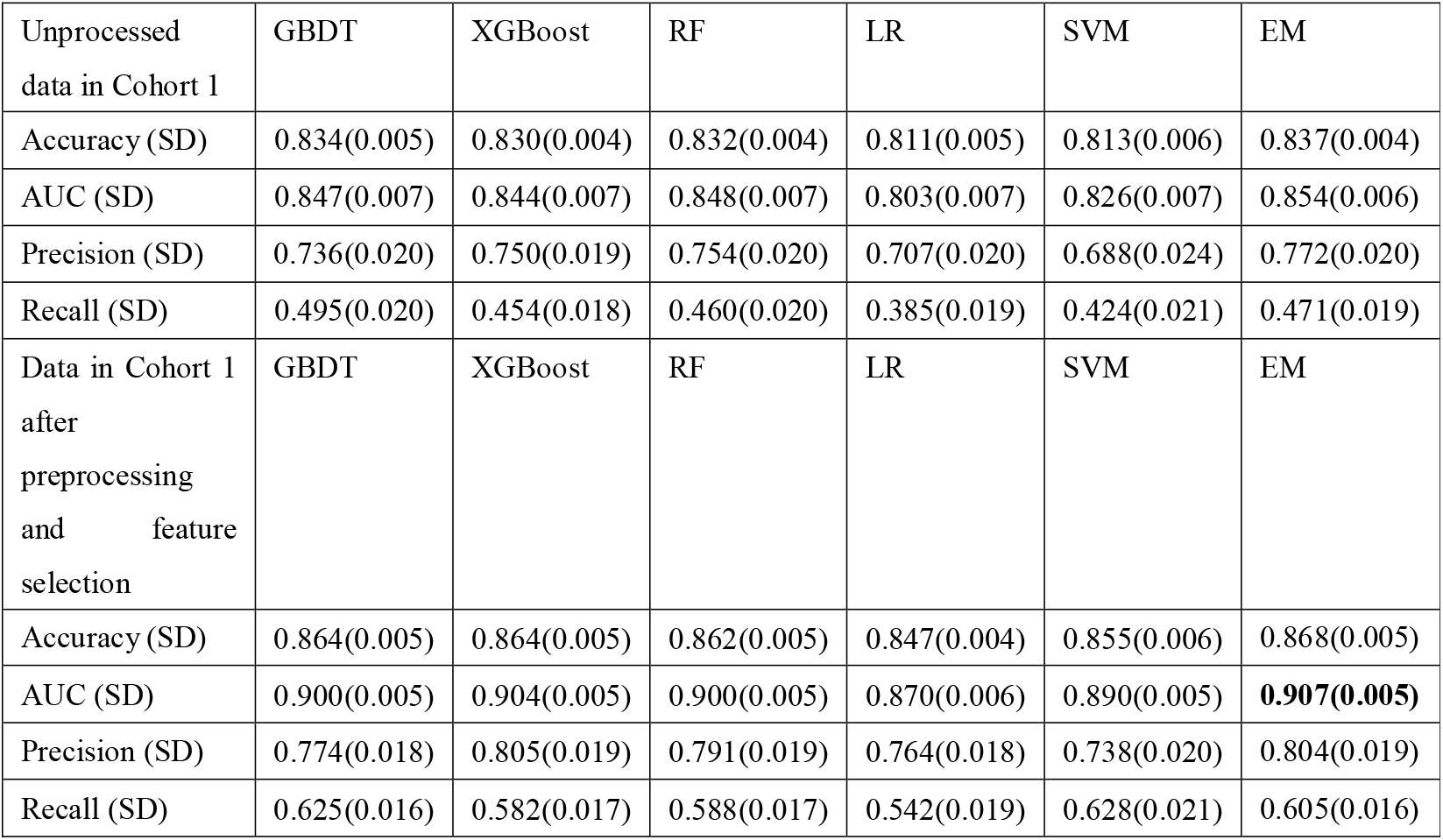
Performance results of different models on Cohort 1

We calculated the mean value of the prediction results of the EM for each patient in 100 rounds to study the changes in precision and recall of EM when the threshold changed from 0 to 1 (Figure 2). With the increasing of the threshold, the precision had a trend of rapidly increasing at first and then slowly increasing, and correspondingly, the recall had a trend of slowly decreasing at first and then rapidly declining. Our goal was to find a reasonable range of thresholds in which the precision and recall can have a practical value. We selected the threshold (0.24) when the recall reached 0.8 and the threshold (0.46) when the precision reached 0.8, and marked it with a dashed line in Figure 2. The precision and recall under these thresholds were 0.656998 and 0.63676, respectively.

**Figure 2.**
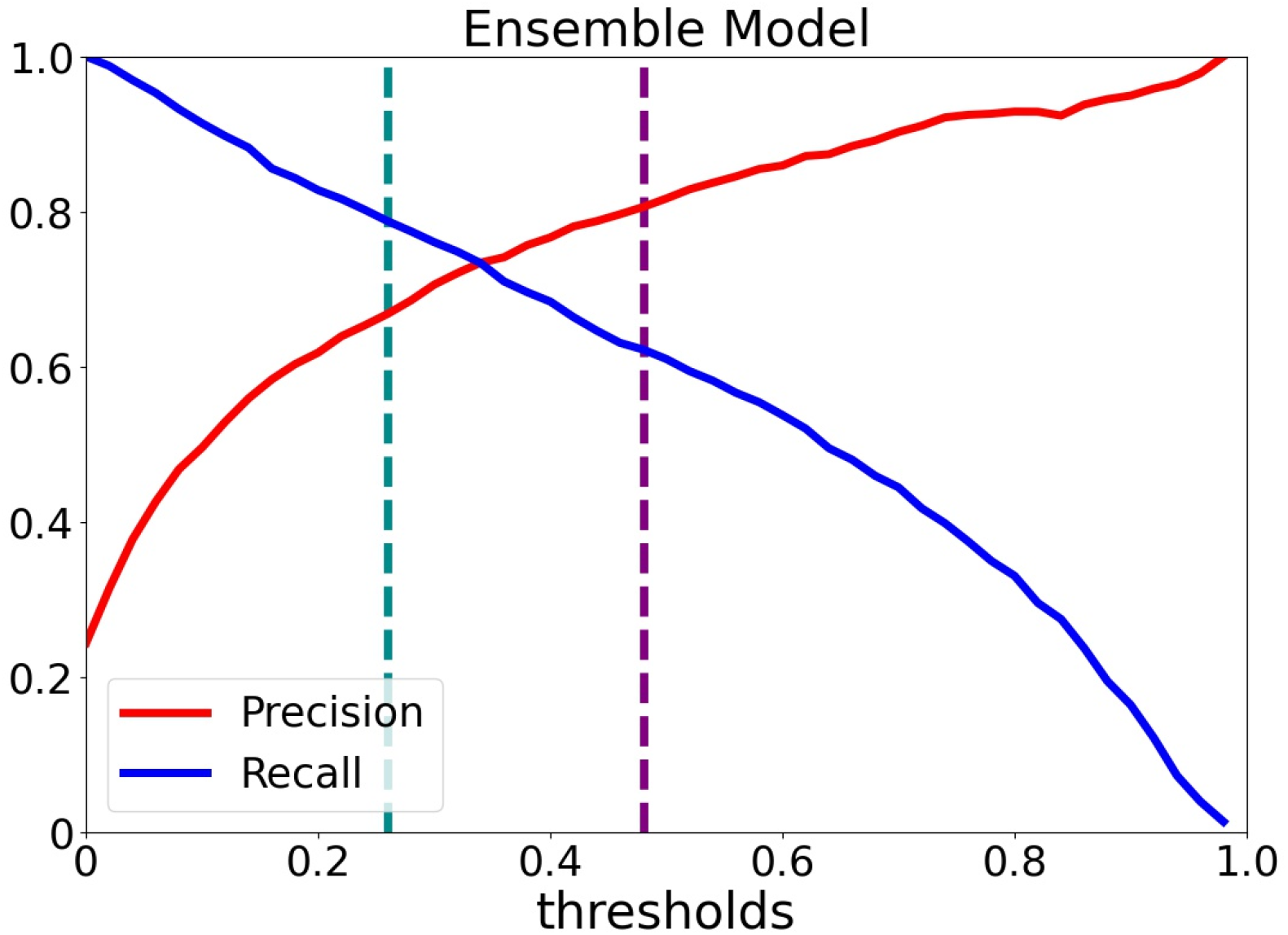
Changes in precision and recall of ensemble model under different thresholds. The x axis represents the threshold, and the y axis represents the value of precision and recall. Precision and recall are represented by red curve and blue curve respectively. The threshold when the precision is 0.8 and the threshold when the recall is 0.8 are indicated by the dashed purple line and the dashed cyan line respectively.

To assess the impact of each feature on mortality in patients, we calculated the mean value of the importance of each feature in the XGBoost over 100 rounds of predictions (Figure 3). MAP, IL-6, and Procalcitonin contributed the most to the decision of XGBoost predictive model. Other features such as Ddimer, age, and CrctProtein also played an important role in the prediction model.

**Figure 3.**
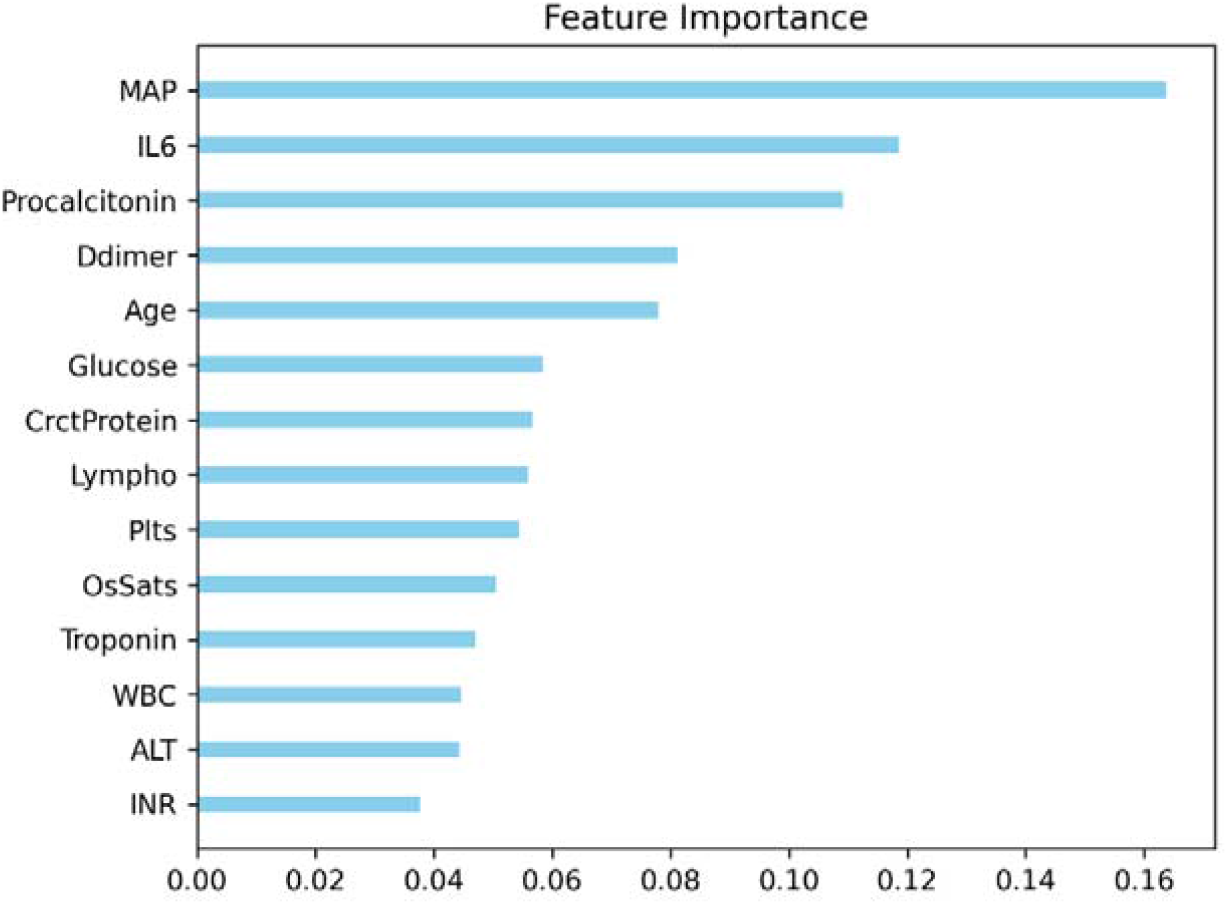
The mean of the importance of each feature in the XGBoost predictive model, over 100 rounds of predictions.

We further explored whether a single clinical feature could be used to stratify patients for mortality risk. To do so, we selected the first six clinical features which have a higher importance. Patients were divided into ten groups according to the value range of each clinical feature, and the number of patients in each group was approximately equal. Patients which procalcitonin is from 0.099 to 0.1 were one group because interval value is too less. The mortality rate in cohort 1 was 0.244 (1148/4711). The mortality rate of each group for six clinical features is shown in Figure 4. From Figure 4, we can see that when these clinical features: MAP < 79.67 mmHg, IL-6 > 72.8 pg/ml, procalcitinon > 0.5 ng/ml, Ddimer > 2.4 mg/ml, age > 69 years, and Glucose > 174.0 mg/dL, patients have higher death risk.

**Figure 4.**
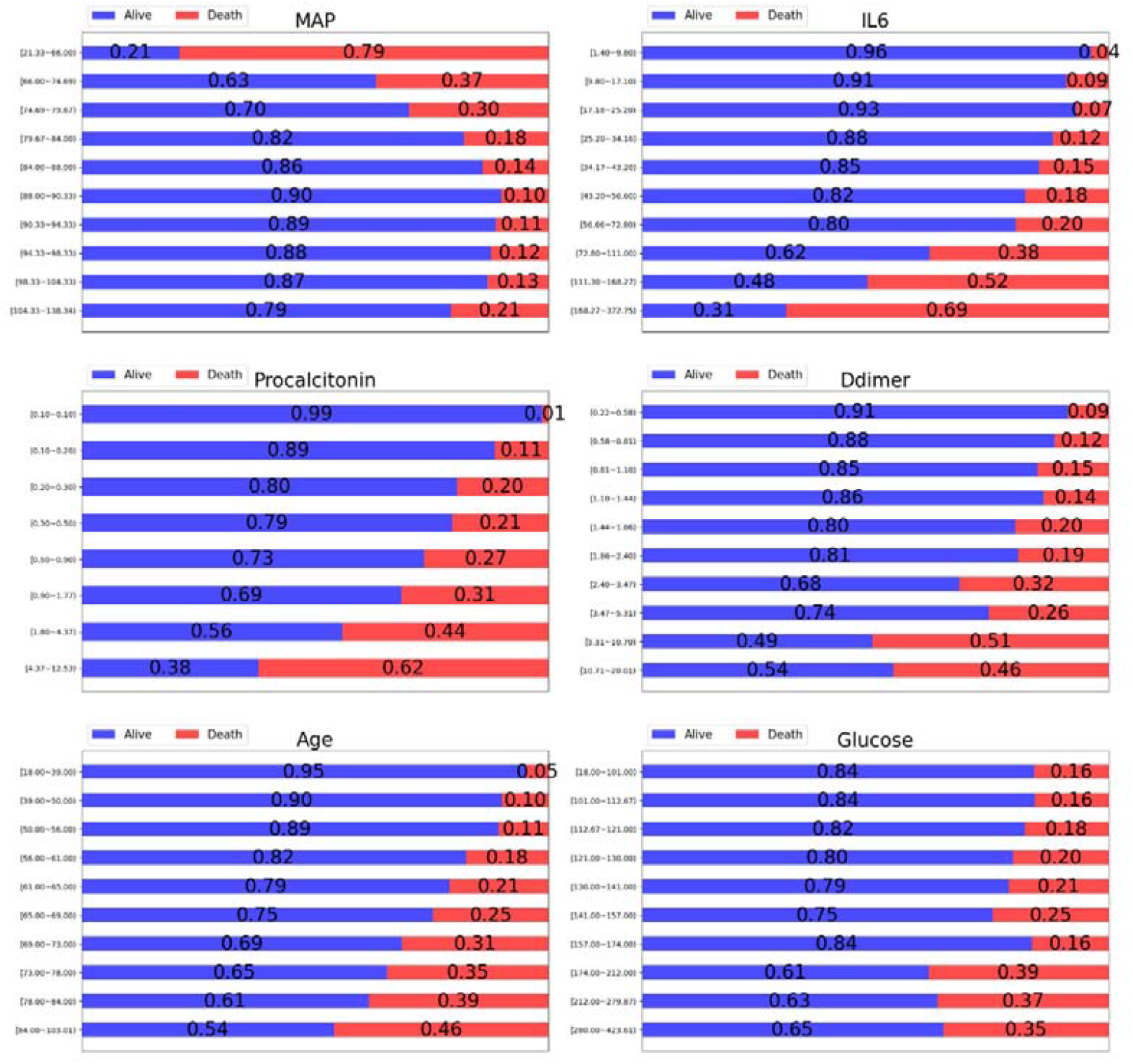
The proportion of patients alive or dead at different values for a single clinical feature. Patients who are alive are shown in blue and those who are dead are shown in red, and the title of each subgraph represents the clinical feature to which it belongs.

### Experimental results on an independent dataset

Finally, we further validate our predictive models on an independent cohort (Cohort 2). First, we selected a set of features (Supplementary Table 6), and then compare the performance of the predictive models in the corresponding subsets of the two cohorts, as described in the methods and materials section. The results of our predictive model on two subsets are shown in Table 6. In general, these predictive models still performed well in a large data set with similar features. EM performs best in three of the four performance metrics, which indicates that EM has a good robustness.

**Table 5.** The percentage of patients who survived or died in the low, moderate, or high-risk group

| Predictive Model | The percentage of patients who survived |  |  | The percentage of patients who died |  |  |
| --- | --- | --- | --- | --- | --- | --- |
|  | Low risk | Moderate risk | High risk | Low risk | Moderate risk | High risk |
| CSS | 88.25 | 60.98 | 22.03 | 11.75 | 39.02 | 77.97 |
| EM | **90.15** | 40.09 | 9.73 | 9.85 | 59.91 | **90.27** |

**Table 6.** Performance of different models on an independent dataset

| Subset 1 | GBDT | XGBoost | RF | LR | SVM | EM |
| --- | --- | --- | --- | --- | --- | --- |
| Accuracy (SD) | 0.850(0.006) | 0.849(0.006) | 0.850(0.006) | 0.828(0.006) | 0.838(0.006) | 0.854(0.005) |
| AUC (SD) | 0.884(0.007) | 0.888(0.007) | 0.887(0.007) | 0.846(0.008) | 0.875(0.007) | 0.893(0.007) |
| Precision (SD) | 0.764(0.020) | 0.797(0.021) | 0.788(0.021) | 0.749(0.020) | 0.723(0.021) | 0.799(0.020) |
| Recall (SD) | 0.616(0.022) | 0.565(0.023) | 0.583(0.023) | 0.515(0.020) | 0.616(0.025) | 0.588(0.022) |
| Subset 2 | GBDT | XGBoost | RF | LR | SVM | EM |
| Accuracy (SD) | 0.800(0.009) | 0.804(0.009) | 0.803(0.009) | 0.804(0.009) | 0.786(0.009) | 0.810(0.009) |
| AUC (SD) | 0.858(0.009) | 0.863(0.010) | 0.860(0.009) | 0.862(0.009) | 0.847(0.009) | 0.871(0.008) |
| Precision (SD) | 0.644(0.023) | 0.687(0.029) | 0.662(0.025) | 0.654(0.024) | 0.612(0.024) | 0.684(0.026) |
| Recall (SD) | 0.533(0.030) | 0.461(0.029) | 0.507(0.031) | 0.533(0.027) | 0.500(0.033) | 0.512(0.028) |

### Comparison with COVID-19 severity scores

Altschul et al. collected COVID-19 patients’ clinical information (Cohort 1) and also developed a predictive model (i.e., COVID-19 severity scores (CSS)) [14]. Therefore, it is possible to direct compare EM and the CSS. In CSS, patients were classified into low risk (0-3 points), moderate risk (4-7 points), and high risk (> 7 points) groups. For the sake of comparison, we also classified patients as low risk group (< 0.4), moderate risk group (0.4-0.7), and high-risk group (> 0.7) according to the average prediction probability of each patient. The comparison results are shown in Table 5. The analysis showed that EM was much better than the CSS: EM was able to assign a higher proportion of patients who survived to the low-risk group and a higher proportion of patients who died to the high-risk group. These results indicated that feature preprocessing, feature selection and EM are helpful in improving predictive performance.

## Discussion

In this study, we developed a novel data preprocessing method to deal with complex clinical data, and an EM to predict high-risk COVID-19 patients. Most importantly, we trained and tested the models in a large cohort and successfully validated the predictive models in a large independent cohort. This is the first study to show that high-risk COVID-19 predictive models and key features are able to reproducible for the predictions in a large independent cohort, suggesting that they could be potentially useful in clinical settings.

By comparing of our model with COVID-19 severity scores [14], we showed that our missing clinical features imputation method and the EM more accurately help physicians predict patients’ mortality risk. In addition, we used GA to select the most appropriate 14 features from the 20 clinical features, and demonstrated that a removing of redundant features significantly improved the performance of the predictive models. The feature importance analysis showed that mean arterial pressure (MAP), interleukin-6 (IL-6), procalcitonin, D-dimer (Ddimer), age, and glucose were the most important features affecting the mortality risk of patients.

We used GA to find the optimal combination coefficient of comprehensive usage of five predictive models to construct the EM. In 100 rounds of half-half cross-validation, the EM achieved the best performance in multiple evaluation indicators. Moreover, we analyzed the precision and recall of each model under different thresholds to help clinicians in making choices according to the availability of clinical information. If physiological indicators, especially clinical features that reflect inflammation, hepatorenal function, and cardiovascular function can be obtained during the patient’s stay in hospital, our models could be easily used to predict high-risk patients timely.

Our study showed that clinical features, such as age, MAP, and features that are associated with physiological status of the patient, can contribute to the predictive model of mortality stratification for COVID-19 in patients. The physiological status of coagulation function (related feature: Ddimer), hepatorenal function (related feature: glucose), and cardiac function (related feature: troponin) also had a noteworthy effect on mortality, which is consistent with previous findings [9-12]. In addition, we provided reference ranges for clinical features to help physicians quickly stratify patients using our models.

The experiments on the Cohort 2 demonstrated the correctness of our feature selection and the robustness of the predictive model. Despite the differences in population characteristics such as age distribution, ethnic proportion between cohort 1 and Cohort 2 (Supplementary Tables 1 and 2), and the inconsistent clinical features adopted (Supplementary Table 3), our prediction method still achieved good performance. These results further confirm that age, MAP, and clinical features related to inflammation, coagulation, hepatorenal function, and cardiovascular function can be used to predict the risk of death in patients with COVID-19.

Finally, we compared our model with those published by others [7, 13, 14, 32-35] (Supplementary Table 7). We first compared features of different models. Overall, age, features associated with inflammation, kidney function, cardiovascular function, and lung function were selected for multiple studies, suggesting that the features we selected were more reasonable. Moreover, we employ more efficient feature selection methods to improve model prediction performance. Then, we compare the frameworks used by different studies. Three studies adopted the gradient boosting framework [7, 33, 34], another three adopted the deep learning framework [13, 32, 35], and one invented a scoring method [14]. Our model (EM) takes advantage of the gradient boosting frameworks (XGBoost and GBDT) with proven predictive performance, as well as the random forest model, logistic regression model, and support vector machine. Finally, we compared performance of different models. Three of the models were not completely consistent with our model in the selection of predicted clinical outcomes, such as severity of the disease[13, 35] and distinguishing COVID-19 patients from other pneumonia patients[32]. Furthermore, our model still achieved a good performance from the perspective of the comparison of the models’ discriminative ability. For the remaining models, our model had the best discriminative performance compared with Rechtman et al.’s model [33] and the COVID-19 severity score[14]. Compared to Barda et al. ‘s study[34] (Supplementary Table 8), our subjects had a higher percentage of deaths and the AUC of our model was slightly lower than their model, but we achieved a higher precision when we achieved the same recall. Compared with the research of Yan et al.[7], their ‘single-tree XGBoost’ model has an outstanding predictive performance (AUC: 0.9506), but they chose only three features and their study cohort consisted of only 485 patients, making their model unreliable and not performing well on the tests of others [8]. In general, our predictive model (EM) is effective in predicting COVID-19 mortality risk.

There are also some limitations in this study. First of all, for cohort 1 (the training set), the patient population we studied was mainly hospitalized patients, and they generally exhibited more severe symptoms and therefore had a higher mortality rate than the general population, which may have caused some bias in our predictive model in the general population. Second, the characteristics of the cohort may vary performance of models and its ability to be validated. For example, the model’s performance was slightly lower in cohort 2 than cohort 1, because the structure of the two cohorts, such as age distribution, sex ratio, mortality rate, etc. is different. In addition, although we adopt functionally similar features, the differences between these features may also be responsible for the difference in model performance between cohorts. Moreover, since most of the clinical features adopted in this study were missing to varying degrees, the imputed data were affected by other data, which may affect the accuracy of the predictive model. Finally, COVID-19 pandemics are often accompanied by surges in patient numbers, resulting in difficulties in collecting all the required clinical features data, which will limit the application of our predictive model.

## Conclusions

In summary, we selected 14 clinical features from 20 clinical features, and comprehensively utilized five predictive models to construct our predictive model: the EM, which had the best performance on multiple predictive evaluation indicators for COVID-19 mortality risk. Most importantly, EM was successfully validated in an independent cohort containing a large number of patients. We also studied the changes of precision and recall of each model under different thresholds, so as to provide reference for doctors to select appropriate thresholds according to medical resources. In addition, feature importance analysis showed that clinical features related to inflammation, hepatorenal function, and cardiovascular function were good predictors for COVID-19 mortality risk, which was consistent with previous studies.

## Data Availability

All data produced in the present work are contained in the manuscript

https://figshare.com/s/79827c396af7df42b3d7.

https://www.ukbiobank.ac.uk/

## List of Abbreviations

OsSats: oxygen saturation
Temp: temperature
MAP: mean arterial pressure
Ddimer: D-dimer
Plts: platelets
INR: international normalized ratio
BUN: blood urea nitrogen
AST: aspartate aminotransferase
ALT: alanine aminotransferase
WBC: white blood cells
Lympho: lymphocytes
IL-6: interleukin-6
CrctProtein: C-reactive protein
KNN: k-nearest neighbor method
GBDT: Gradient Boosted Decision Tree
XGBoost: Extreme Gradient Boosting
RF: Random Forest
LR: Logistic Regression
SVM: Support Vector Machine
EM: Ensemble Model
ROC: Receiver Operating Characteristic
AUC: Area Under ROC Curve
TP: True Positive
FP: False Positive, TN: True Negative
FN: False Negative
CSS: COVID-19 severity scores

## Declarations

### Ethics approval and consent to participate

Informed consent was waived due to the nature of study being retrospective.

### Consent for publication

Not applicable

### Availability of Data and materials

All data after de-identification will be made available with publication upon request to the corresponding author. The source code for data analysis is available.

## Authors’ contributions

JL and EW were responsible for the conception and design of the study. YW provided support. JL, XL and EW were responsible for the implementation and analysis of the algorithm. JH, MA, JL, XL and ED were responsible for data collection. JH and MA were responsible for model validation on cohort 2. XL, JL and EW were responsible for manuscript writing.

## Acknowledgements

Not applicable

## Reference

1. Arabi YM, Murthy S, Webb S. COVID-19: a novel coronavirus and a novel challenge for critical care. Intensive Care Medicine. 2020;46(5):833–6.

2. Grasselli G, Pesenti A, Cecconi M. Critical Care Utilization for the COVID-19 Outbreak in Lombardy, Italy: Early Experience and Forecast During an Emergency Response. JAMA. 2020;323(16):1545–6.

3. Xie J, Tong Z, Guan X, Du B, Qiu H, Slutsky AS. Critical care crisis and some recommendations during the COVID-19 epidemic in China. Intensive Care Medicine. 2020;46(5):837–40.

4. Guan W-j, Ni Z-y, Hu Y, Liang W-h, Ou C-q, He J-x, et al. Clinical Characteristics of Coronavirus Disease 2019 in China. New England Journal of Medicine. 2020;382(18):1708–20.

5. Wu Z, McGoogan JM. Characteristics of and Important Lessons From the Coronavirus Disease 2019 (COVID-19) Outbreak in China: Summary of a Report of 72lll314 Cases From the Chinese Center for Disease Control and Prevention. JAMA. 2020;323(13):1239–42.

6. Ji Y, Ma Z, Peppelenbosch MP, Pan Q. Potential association between COVID-19 mortality and health-care resource availability. The Lancet Global Health. 2020;8(4):e480.

7. Yan L, Zhang H-T, Goncalves J, Xiao Y, Wang M, Guo Y, et al. An interpretable mortality prediction model for COVID-19 patients. Nature Machine Intelligence. 2020;2(5):283–8.

8. Barish M, Bolourani S, Lau LF, Shah S, Zanos TP. External validation demonstrates limited clinical utility of the interpretable mortality prediction model for patients with COVID-19. Nature Machine Intelligence. 2021;3(1):25–7.

9. Yang X, Yu Y, Xu J, Shu H, Xia Ja, Liu H, et al. Clinical course and outcomes of critically ill patients with SARS-CoV-2 pneumonia in Wuhan, China: a single-centered, retrospective, observational study. The Lancet Respiratory Medicine. 2020;8(5):475–81.

10. Richardson S, Hirsch JS, Narasimhan M, Crawford JM, McGinn T, Davidson KW, et al. Presenting Characteristics, Comorbidities, and Outcomes Among 5700 Patients Hospitalized With COVID-19 in the New York City Area. JAMA. 2020;323(20):2052–9.

11. Cheng Y, Luo R, Wang K, Zhang M, Wang Z, Dong L, et al. Kidney disease is associated with in-hospital death of patients with COVID-19. Kidney International. 2020;97(5):829–38.

12. Wu S, Du Z, Shen S, Zhang B, Yang H, Li X, et al. Identification and Validation of a Novel Clinical Signature to Predict the Prognosis in Confirmed Coronavirus Disease 2019 Patients. Clinical Infectious Diseases. 2020;71(12):3154–62.

13. Liang W, Yao J, Chen A, Lv Q, Zanin M, Liu J, et al. Early triage of critically ill COVID-19 patients using deep learning. Nature Communications. 2020;11(1):3543.

14. Altschul DJ, Unda SR, Benton J, de la Garza Ramos R, Cezayirli P, Mehler M, et al. A novel severity score to predict inpatient mortality in COVID-19 patients. Scientific Reports. 2020;10(1):16726.

15. Wynants L, Van Calster B, Collins GS, Riley RD, Heinze G, Schuit E, et al. Prediction models for diagnosis and prognosis of covid-19: systematic review and critical appraisal. BMJ. 2020;369:m1328.

16. Chen T, Guestrin C. XGBoost: A Scalable Tree Boosting System. Proceedings of the 22nd ACM SIGKDD International Conference on Knowledge Discovery and Data Mining; San Francisco, California, USA: Association for Computing Machinery; 2016. p. 785–94.

17. Sudlow C, Gallacher J, Allen N, Beral V, Burton P, Danesh J, et al. UK biobank: an open access resource for identifying the causes of a wide range of complex diseases of middle and old age. PLoS Med. 2015;12(3):e1001779.

18. Barbour V. UK Biobank: a project in search of a protocol? The Lancet. 2003;361(9370):1734–8.

19. Nath KA, Grande JP, Farrugia G, Croatt AJ, Belcher JD, Hebbel RP, et al. Age sensitizes the kidney to heme protein-induced acute kidney injury. American Journal of Physiology-Renal Physiology. 2013;304(3):F317–F25.

20. Ybarra J, Fernández S, Sánchez-Hernández J, Romeo JH, Ballesta-Lopez C, Guell J, et al. Serum alanine aminotransferase predicts interventricular septum thickness and left ventricular mass in patients with nonalcoholic fatty liver disease. European Journal of Gastroenterology & Hepatology. 2014;26(6):654–60.

21. Palekar NA, Naus R, Larson SP, Ward J, Harrison SA. Clinical model for distinguishing nonalcoholic steatohepatitis from simple steatosis in patients with nonalcoholic fatty liver disease. Liver International. 2006;26(2):151–6.

22. Cano N. Bench-to-bedside review: Glucose production from the kidney. Critical Care. 2002;6(4):317.

23. McNabb WR, Noormohamed FH, Lant AF. The Effects of Enalapril on Blood Pressure and the Kidney in Normotensive Subjects under Altered Sodium Balance. Journal of Hypertension. 1986;4(1):39–47.

24. Frasca D, Blomberg BB. Inflammaging decreases adaptive and innate immune responses in mice and humans. Biogerontology. 2016;17(1):7–19.

25. Galetto-Lacour A, Zamora SA, Gervaix A. Bedside Procalcitonin and C-Reactive Protein Tests in Children With Fever Without Localizing Signs of Infection Seen in a Referral Center. Pediatrics. 2003;112(5):1054.

26. Kennergren C, Mantovani V, Lönnroth P, Nyström B, Berglin E, Hamberger A. Monitoring of Extracellular Aspartate Aminotransferase and Troponin T by Microdialysis during and after Cardioplegic Heart Arrest. Cardiology. 1999;92(3):162–70.

27. Schindhelm RK, Dekker JM, Nijpels G, Bouter LM, Stehouwer CDA, Heine RJ, et al. Alanine aminotransferase predicts coronary heart disease events: A 10-year follow-up of the Hoorn Study. Atherosclerosis. 2007;191(2):391–6.

28. Verni CC, Davila A, Jr, Sims CA, Diamond SL. D-Dimer and Fibrin Degradation Products Impair Platelet Signaling: Plasma D-Dimer Is a Predictor and Mediator of Platelet Dysfunction During Trauma. The Journal of Applied Laboratory Medicine. 2020;5(6):1253–64.

29. Ellis RJ, Mayo MS, Bodensteiner DM. Ciprofloxacin–warfarin coagulopathy: A case series. American Journal of Hematology. 2000;63(1):28–31.

30. Yang J, Honavar V. Feature Subset Selection Using a Genetic Algorithm. In: Liu H, Motoda H, editors. Feature Extraction, Construction and Selection: A Data Mining Perspective. Boston, MA: Springer US; 1998. p. 117–36.

31. Blickle T, Thiele L. A Comparison of Selection Schemes Used in Evolutionary Algorithms. Evolutionary Computation. 1996;4(4):361–94.

32. Wang S, Zha Y, Li W, Wu Q, Li X, Niu M, et al. A fully automatic deep learning system for COVID-19 diagnostic and prognostic analysis. European Respiratory Journal. 2020;56(2):2000775.

33. Rechtman E, Curtin P, Navarro E, Nirenberg S, Horton MK. Vital signs assessed in initial clinical encounters predict COVID-19 mortality in an NYC hospital system. Scientific Reports. 2020;10(1):21545.

34. Barda N, Riesel D, Akriv A, Levy J, Finkel U, Yona G, et al. Developing a COVID-19 mortality risk prediction model when individual-level data are not available. Nature Communications. 2020;11(1):4439.

35. Ning W, Lei S, Yang J, Cao Y, Jiang P, Yang Q, et al. Open resource of clinical data from patients with pneumonia for the prediction of COVID-19 outcomes via deep learning. Nature Biomedical Engineering. 2020;4(12):1197–207.

