## Supplementary Tables and Figures for "An ensemble prediction model for COVID-19 mortality risk"

**Supplementary Table 1. Clinical information statistics in cohort 1.**

| **Clinical Information** | **Mean** | **Std** | **Min** | **25%** | **50%** | **75%** | **Max** | **NA count** | **NA percentage (%)** |
| --- | --- | --- | --- | --- | --- | --- | --- | --- | --- |
| Age (years) | 63.36956 | 16.70179 | 18 | 54 | 65 | 76 | 103 | 0 | 0 |
| Oxygen saturation (%) | 92.88644 | 8.18966 | 11 | 91 | 95 | 98 | 100 | 167 | 3.544894927 |
| Temperature (°C) | 37.01411 | 4.171799 | -17.7778 | 36.72222 | 37.11111 | 37.72222 | 50 | 153 | 3.247718107 |
| Mean Arterial Pressure (mmHg) | 85.79015 | 16.80901 | 21.33333 | 77.66667 | 87.33333 | 96.66667 | 138.3333 | 222 | 4.712375292 |
| D-dimer (mg/ml) | 4.205255 | 5.665569 | 0.22 | 0.9 | 1.83 | 3.94 | 20.00001 | 1101 | 23.37083422 |
| Platelets (k/mm^3^) | 235.5437 | 107.8714 | 1 | 164.75 | 216 | 282 | 1226 | 183 | 3.884525578 |
| International normalized ratio | 1.236779 | 0.975106 | 0.8 | 1 | 1.1 | 1.2 | 17.0001 | 502 | 10.6559117 |
| Blood urea nitrogen (mg/dL) | 31.20384 | 31.51133 | 4.999 | 12 | 19 | 38 | 301 | 605 | 12.84228402 |
| Creatinine (µmol/L) | 2.029297 | 2.661654 | 0.19999 | 0.8 | 1.1 | 1.88 | 31.66 | 190 | 4.033113989 |
| Sodium (mmol/L) | 138.1303 | 7.531861 | 105 | 134 | 137 | 141 | 170.001 | 236 | 5.009552112 |
| Glucose (mg/L) | 180.3811 | 121.4106 | 18 | 113 | 137 | 198 | 1000.001 | 1373 | 29.1445553 |
| Aspartate aminotransferase (U/L) | 67.58342 | 210.756 | 13 | 27 | 40 | 65 | 10000 | 305 | 6.474209297 |
| Alanine aminotransferase (U/L) | 45.30904 | 111.5789 | 6 | 17 | 27 | 45 | 3228 | 256 | 5.434090427 |
| White Blood Cells (per mm^3^) | 8.8062 | 7.301112 | 0.2 | 5.6 | 7.5 | 10.4 | 219.7 | 179 | 3.799617916 |
| Lymphocytes (per mm^3^) | 1.360371 | 4.951058 | 0.1 | 0.7 | 1 | 1.4 | 209.1 | 179 | 3.799617916 |
| Interleukin-6 (pg/ml) | 283.8747 | 3366.74 | 1.39999 | 16.02 | 38 | 85.25 | 111040 | 2636 | 55.95414986 |
| Ferritin (µg/L) | 1506.678 | 3604.729 | 1.999 | 379.75 | 800.5 | 1638 | 100000 | 1407 | 29.86627043 |
| C-Reactive protein (mg/L) | 12.67108 | 11.17503 | 0.4999 | 4.1 | 9.9 | 18.2 | 100.0001 | 876 | 18.59477818 |
| Procalcitonin (ng/ml) | 2.492415 | 7.706973 | 0.0999 | 0.1 | 0.2 | 1 | 50.0001 | 1714 | 36.38293356 |
| Troponin (ng/ml) | 0.059678 | 0.287668 | 0.00999 | 0.01 | 0.01 | 0.03 | 9.56 | 641 | 13.60645298 |
| Length of hospital stay | 7.160263214 | 7.029781979 | 0 | 3 | 5 | 9 | 56 | 0 | 0 |

Note: Of the 4,711 cases, 466 were white, 1,743 African Americans, 1,753 Hispanics and 121 Asians. The number of female patients is 2200.[1] ; Missing data is represented by NA.

**Supplementary Table 2. Clinical information statistics in cohort 2.**

| **Clinical features** | **mean** | **std** | **min** | **25%** | **50%** | **75%** | **max** | **NA count** | **NA percentage (%)** |
| --- | --- | --- | --- | --- | --- | --- | --- | --- | --- |
| Age (years) | 65.48049 | 8.616102 | 50 | 58 | 65 | 73 | 84 | 0 | 0 |
| Haemoglobin concentration (g/dL) | 14.14331 | 1.309253 | 5.74 | 13.27 | 14.12 | 15.0575 | 20.52 | 764 | 4.838505 |
| Red blood cell count (cells/L) | 4.536164 | 0.434811 | 1.803 | 4.236 | 4.52 | 4.82 | 7.298 | 764 | 4.838505 |
| Haematocrit percentage (%) | 41.02078 | 3.692196 | 15.46 | 38.5 | 41 | 43.59 | 63.46 | 764 | 4.838505 |
| Mean corpuscular volume (pg) | 90.60556 | 4.807659 | 58.4 | 88 | 90.78 | 93.56 | 134.6 | 764 | 4.838505 |
| Mean corpuscular haemoglobin (pg) | 31.25129 | 1.993003 | 17.42 | 30.3 | 31.37 | 32.4 | 65.12 | 764 | 4.838505 |
| Mean corpuscular haemoglobin concentration (g/dL) | 34.48474 | 1.040153 | 24.2 | 33.85 | 34.44 | 35.1 | 57.26 | 764 | 4.838505 |
| Red blood cell distribution width (%) | 13.52138 | 1.052667 | 11.33 | 12.9 | 13.34 | 13.9 | 36.29 | 764 | 4.838505 |
| Nucleated red blood cell count (10^9 cells/L) | 0.002243 | 0.024197 | 0 | 0 | 0 | 0 | 0.82 | 803 | 5.085497 |
| Nucleated red blood cell percentage (%) | 0.037879 | 0.39592 | 0 | 0 | 0 | 0 | 10.91 | 803 | 5.085497 |
| Reticulocyte percentage (%) | 1.399372 | 0.936986 | 0.024 | 0.993 | 1.31 | 1.687 | 49.199 | 1032 | 6.535782 |
| Reticulocyte count (10^12 cells/Litre) | 0.06349 | 0.041003 | 0.001 | 0.044 | 0.059 | 0.077 | 2.045 | 1032 | 6.535782 |
| Mean reticulocyte volume (fL) | 106.0377 | 8.113087 | 52.92 | 101.4 | 106.055 | 110.9375 | 182.28 | 1032 | 6.535782 |
| Mean sphered cell volume (fL) | 82.65293 | 5.483405 | 46.07 | 79.02 | 82.44 | 86 | 121.45 | 1041 | 6.59278 |
| Immature reticulocyte fraction (ratio) | 0.293926 | 0.062167 | 0 | 0.252 | 0.292 | 0.335 | 0.644 | 1041 | 6.59278 |
| High light scatter reticulocyte percentage (%) | 0.420558 | 0.232918 | 0.003 | 0.263 | 0.38 | 0.53 | 7.882 | 1041 | 6.59278 |
| High light scatter reticulocyte count (10^12 cells/L) | 0.019084 | 0.010581 | 0 | 0.012 | 0.017 | 0.024 | 0.33 | 1041 | 6.59278 |
| Diastolic blood pressure (mmHg) | 82.02513 | 10.50363 | 30 | 75 | 82 | 89 | 134 | 390 | 2.469918 |
| Systolic blood pressure (mmHg) | 134.1949 | 18.33459 | 82 | 121 | 132 | 145 | 254 | 390 | 2.469918 |
| White blood cell count (10^9 cells/L) | 6.963708 | 2.095475 | 0.1 | 5.7 | 6.73 | 7.99 | 127.5 | 765 | 4.844839 |
| Lymphocyte count (10^9 cells/L) | 1.995828 | 1.22414 | 0.03 | 1.54 | 1.9 | 2.31 | 118.57 | 793 | 5.022166 |
| Neutrophill count (10^9 cells/L) | 4.269461 | 1.432635 | 0.06 | 3.3 | 4.09 | 5.03 | 14.84 | 793 | 5.022166 |
| Eosinophill count (10^9 cells/L) | 0.179466 | 0.141789 | 0 | 0.1 | 0.14 | 0.22 | 3.4 | 793 | 5.022166 |
| Basophill count (10^9 cells/L) | 0.035706 | 0.048384 | 0 | 0.01 | 0.02 | 0.05 | 1.43 | 793 | 5.022166 |
| Lymphocyte percentage (%) | 29.04088 | 7.569034 | 1.01 | 24.1 | 28.6 | 33.61 | 93 | 792 | 5.015833 |
| Monocyte percentage (%) | 7.001157 | 2.982392 | 0 | 5.47 | 6.75 | 8.2 | 75.6 | 792 | 5.015833 |
| Neutrophill percentage (%) | 60.77478 | 8.66898 | 1.6 | 55.5 | 61.1 | 66.4 | 94.5 | 792 | 5.015833 |
| Eosinophill percentage (%) | 2.610865 | 1.946774 | 0 | 1.4 | 2.17 | 3.3 | 77.9 | 792 | 5.015833 |
| Basophill percentage (%) | 0.572766 | 0.553077 | 0 | 0.3 | 0.43 | 0.68 | 13.41 | 792 | 5.015833 |
| Platelet count (10^9 cells/L) | 252.1626 | 60.88152 | 14.1 | 212 | 246.2 | 286.9 | 967 | 765 | 4.844839 |
| Platelet crit (%) | 0.232781 | 0.050096 | 0.014 | 0.2 | 0.229 | 0.261 | 0.811 | 774 | 4.901837 |
| Mean platelet volume (fL) | 9.35372 | 1.072022 | 6.25 | 8.6 | 9.24 | 9.97 | 14.9 | 765 | 4.844839 |
| Platelet distribution width (%) | 16.50215 | 0.533886 | 14.36 | 16.12 | 16.43 | 16.8 | 19.52 | 774 | 4.901837 |
| Creatinine (enzymatic) in urine (μmol/L) | 9616.533 | 6167.608 | 622 | 4835.75 | 8362.5 | 13055 | 67604 | 634 | 4.015199 |
| Sodium in urine (mmol/L) | 84.33877 | 46.58997 | 10 | 47.6 | 76.2 | 112.825 | 323.2 | 666 | 4.217859 |
| Pulse rate (bpm) | 69.12864 | 11.35972 | 34 | 61 | 68 | 76 | 150 | 390 | 2.469918 |
| Body fat percentage (%) | 32.13543 | 8.535141 | 5.3 | 25.8 | 31.6 | 38.5 | 59.9 | 356 | 2.254592 |
| Whole body fat mass (Kg) | 26.29037 | 10.23083 | 5 | 19.3 | 24.5 | 31.6 | 99.7 | 365 | 2.31159 |
| Whole body fat-free mass (Kg) | 54.44017 | 11.83243 | 29.5 | 44.3 | 52.1 | 63.8 | 99.9 | 350 | 2.216593 |
| Whole body water mass (Kg) | 39.8454 | 8.665677 | 21.6 | 32.4 | 38.1 | 46.6 | 77 | 348 | 2.203927 |
| Body mass index (Kg/m^2) | 28.3288 | 5.110593 | 15.3 | 24.8 | 27.6 | 31 | 62 | 348 | 2.203927 |
| Basal metabolic rate (KJ) | 6789.843 | 1420.368 | 3833 | 5615 | 6561 | 7799 | 14192 | 348 | 2.203927 |
| Trunk fat percentage (%) | 31.92367 | 7.909053 | 3 | 26.7 | 31.8 | 37.3 | 61.9 | 358 | 2.267258 |
| Trunk fat mass (Kg) | 14.49804 | 5.414035 | 0.7 | 10.7 | 13.9 | 17.6 | 49.1 | 359 | 2.273591 |
| Trunk fat-free mass (kg) | 30.12582 | 6.07396 | 14.2 | 24.9 | 29 | 35 | 53.7 | 361 | 2.286257 |
| Trunk predicted mass (kg) | 28.88888 | 5.891231 | 13.6 | 23.8 | 27.75 | 33.7 | 51.6 | 362 | 2.29259 |

Note: The ethnic backgrounds of the 15,790 patients were as follows: British (13177), any other white background (469), Irish (396), Indian (388), other ethnic group (251), Caribbean (250), African (238), Pakistani (197), any other Asian background (98), prefer not to answer (49), Chinese (41), any other mixed background (41), White and Black Caribbean (33), White and Asian (30), Bangladeshi (25), White and Black African (25), White (21), Do not know (11), any other Black background (8), Black or Black British (3), Mixed (3). The number of female patients is 8342.; Missing data is represented by NA.

**Supplementary Table 3. Functionally related features**

| **Type of features** | **The original features** | **Functionally related features** | **Details of the features** |
| --- | --- | --- | --- |
| Age | 1. Age | 1. Age | COVID19_First_Test_Date |
|  |  |  | 34: Year of birth |
| Pulmonary function & hemoglobin | 2. Oxygen saturation | 2. Red blood cell count | 30010: Red blood cell (erythrocyte) count |
|  |  | 3. Haemoglobin concentration | 30020: Haemoglobin concentration |
|  |  | 4. Haematocrit percentage | 30030: Haematocrit percentage |
|  |  | 5. Mean corpuscular volume | 30040: Mean corpuscular volume |
|  |  | 6. Mean corpuscular haemoglobin | 30050: Mean corpuscular haemoglobin |
|  |  | 7. Mean corpuscular haemoglobin concentration | 30060: Mean corpuscular haemoglobin concentration |
|  |  | 8. Red blood cell distribution width | 30070: Red blood cell (erythrocyte) distribution width |
|  |  | 9. Nucleated red blood cell count | 30170: Nucleated red blood cell count |
|  |  | 10. Nucleated red blood cell percentage | 30230: Nucleated red blood cell percentage |
|  |  | 11. Reticulocyte percentage | 30240: Reticulocyte percentage |
|  |  | 12. Reticulocyte count | 30250: Reticulocyte count |
|  |  | 13. Mean reticulocyte volume | 30260: Mean reticulocyte volume |
|  |  | 14. Mean sphered cell volume | 30270: Mean sphered cell volume |
|  |  | 15. Immature reticulocyte fraction | 30280: Immature reticulocyte fraction |
|  |  | 16. High light scatter reticulocyte percentage | 30290: High light scatter reticulocyte percentage |
|  |  | 17. High light scatter reticulocyte count | 30300: High light scatter reticulocyte count |
|  |  | 18. Number of lung diseases | Illness:ICD: Diseases of the respiratory system |
|  |  |  | Illness:J22 Unspecified acute lower respiratory infection |
|  |  |  | Illness:O99.5 Diseases of the respiratory system complicating pregnancy- childbirth and the puerperium |
|  |  |  | Illness:J95.8 Other postprocedural respiratory disorders |
|  |  |  | Illness:J06.9 Acute upper respiratory infection- unspecified |
|  |  |  | Illness:Z87.0 Personal history of diseases of the respiratory system |
|  |  |  | Illness:J44.0 Chronic obstructive pulmonary disease with acute lower respiratory infection |
|  |  |  | Illness:Z82.5 Family history of asthma and other chronic lower respiratory diseases |
|  |  |  | Illness:J96.00 Acute respiratory failure; Type I [hypoxic] |
|  |  |  | Illness:J96.0 Acute respiratory failure |
|  |  |  | Illness:Z85.2 Personal history of malignant neoplasm of other respiratory and intrathoracic organs |
|  |  |  | Illness:R09.8 Other specified symptoms and signs involving the circulatory and respiratory systems |
|  |  |  | Illness:J10.1 Influenza with other respiratory manifestations- influenza virus identified |
|  |  |  | Illness:Z11.1 Special screening examination for respiratory tuberculosis |
|  |  |  | Illness:Z99.1 Dependence on respirator |
|  |  |  | Illness:J80 Adult respiratory distress syndrome |
|  |  |  | Illness:J96.11 Chronic respiratory failure; Type II [hypercapnic] |
|  |  |  | Illness:J11.1 Influenza with other respiratory manifestations- virus not identified |
|  |  |  | Illness:J96.1 Chronic respiratory failure |
|  |  |  | Illness:T17.9 Foreign body in respiratory tract- part unspecified |
|  |  |  | Illness:J39.8 Other specified diseases of upper respiratory tract |
|  |  |  | Illness:R04.8 Haemorrhage from other sites in respiratory passages |
|  |  |  | Illness:J98.8 Other specified respiratory disorders |
|  |  |  | Illness:J96.01 Acute respiratory failure; Type II [hypercapnic] |
|  |  |  | Illness:Z12.2 Special screening examination for neoplasm of respiratory organs |
|  |  |  | Illness:B90.9 Sequelae of respiratory and unspecified tuberculosis |
|  |  |  | Illness:U04.9 Severe acute respiratory syndrome [SARS]- unspecified |
|  |  |  | Illness:T17.8 Foreign body in other and multiple parts of respiratory tract |
|  |  |  | Illness:D38.5 Other respiratory organs |
|  |  |  | Illness:Z83.6 Family history of diseases of the respiratory system |
|  |  |  | Illness:J96.10 Chronic respiratory failure; Type I [hypoxic] |
|  |  |  | Illness:Z80.2 Family history of malignant neoplasm of other respiratory and intrathoracic organs |
|  |  |  | Illness:J68.2 Upper respiratory inflammation due to chemicals- gases- fumes and vapours- not elsewhere classified |
|  |  |  | Illness:Z13.83 Encounter for screening for respiratory disorder NEC |
|  |  |  | Illness:J96.09 Acute respiratory failure; Type unspecified |
|  |  |  | Illness:J96.90 Respiratory failure- unspecified; Type I [hypoxic] |
|  |  |  | Illness:J96.9 Respiratory failure- unspecified |
|  |  |  | Illness:R09.2 Respiratory arrest |
|  |  |  | Illness:J96.91 Respiratory failure unspecified; Type II [hypercapnic] |
|  |  |  | Illness:B97.4 Respiratory synctial virus as the cause of diseases classified to other chapters |
|  |  |  | Illness:J96.99 Respiratory failure- unspecified; Type unspecified |
|  |  |  | Illness:J12.1 Respiratory syncytial virus pneumonia |
|  |  |  | Illness:J98.9 Respiratory disorder- unspecified |
|  |  |  | Illness:J99.1 Respiratory disorders in other diffuse connective tissue disorders |
|  |  |  | Illness:J99.8 Respiratory disorders in other diseases classified elsewhere |
|  |  |  | Illness:A16.9 Respiratory tuberculosis unspecified- without mention of bacteriological or histological confirmation |
|  |  |  | 22130: Doctor diagnosed COPD (chronic obstructive pulmonary disease) |
|  |  |  | 22170: Recent medication for COPD (Chronic Obstructive Pulmonary Disease) |
|  |  |  | 22127: Doctor diagnosed asthma |
|  |  |  | 22167: Recent medication for asthma |
|  |  |  | Illness:Z82.5 Family history of asthma and other chronic lower respiratory diseases |
|  |  |  | Illness:J45.0 Predominantly allergic asthma |
|  |  |  | Illness:J46 Status asthmaticus |
|  |  |  | Illness:J45.1 Nonallergic asthma |
|  |  |  | Illness:Y55.6 Antiasthmatics- not elsewhere classified |
|  |  |  | Illness:J45.8 Mixed asthma |
|  |  |  | Illness:J45.9 Asthma- unspecified |
| Thrombin & thrombus | 3. International normalized ratio (INR)  4. D-dimer  5. Platelet (Plts) | 19. Platelet count | 30080: Platelet count |
|  |  | 20. Platelet crit | 30090: Platelet crit |
|  |  | 21. Mean platelet volume | 30100: Mean platelet (thrombocyte) volume |
|  |  | 22. Platelet distribution width | 30110: Platelet distribution width |
|  |  | 23. Number of thrombotic diseases | Illness:I80.2 Phlebitis and thrombophlebitis of other deep vessels of lower extremities |
|  |  |  | Illness:D69.6 Thrombocytopenia- unspecified |
|  |  |  | Illness:D47.3 Essential (haemorrhagic) thrombocythaemia |
|  |  |  | Illness:I80.0 Phlebitis and thrombophlebitis of superficial vessels of lower extremities |
|  |  |  | Illness:I74.3 Embolism and thrombosis of arteries of the lower extremities |
|  |  |  | Illness:I63.3 Cerebral infarction due to thrombosis of cerebral arteries |
|  |  |  | Illness:I24.0 Coronary thrombosis not resulting in myocardial infarction |
|  |  |  | Illness:I82.8 Embolism and thrombosis of other specified veins |
|  |  |  | Illness:I80.8 Phlebitis and thrombophlebitis of other sites |
|  |  |  | Illness:D68.6 Other Thrombophilia |
|  |  |  | Illness:D69.5 Secondary thrombocytopenia |
|  |  |  | Illness:I80.3 Phlebitis and thrombophlebitis of lower extremities- unspecified |
|  |  |  | Illness:I23.6 Thrombosis of atrium- auricular appendage and ventricle as current complications following acute myocardial infarction |
|  |  |  | Illness:I51.3 Intracardiac thrombosis- not elsewhere classified |
|  |  |  | Illness:D69.3 Idiopathic thrombocytopenic purpura |
|  |  |  | Illness:I74.2 Embolism and thrombosis of arteries of the upper extremities |
|  |  |  | Illness:I74.9 Embolism and thrombosis of unspecified artery |
|  |  |  | Illness:D75.2 Essential thrombocytosis |
|  |  |  | Illness:D68.5 Primary Thrombophilia |
|  |  |  | Illness:I84.0 Internal thrombosed haemorrhoids |
|  |  |  | Illness:I84.3 External thrombosed haemorrhoids |
|  |  |  | Illness:I84.7 Unspecified thrombosed haemorrhoids |
|  |  |  | Illness:I74.1 Embolism and thrombosis of other and unspecified parts of aorta |
|  |  |  | Illness:I80.1 Phlebitis and thrombophlebitis of femoral vein |
|  |  |  | Illness:I82.2 Embolism and thrombosis of vena cava |
|  |  |  | Illness:I80.9 Phlebitis and thrombophlebitis of unspecified site |
|  |  |  | Illness:I74.5 Embolism and thrombosis of iliac artery |
|  |  |  | Illness:D69.4 Other primary thrombocytopenia |
|  |  |  | Illness:Y44.5 Thrombolytic drugs |
|  |  |  | Illness:O87.1 Deep phlebothrombosis in the puerperium |
|  |  |  | Illness:I74.0 Embolism and thrombosis of abdominal aorta |
|  |  |  | Illness:I81 Portal vein thrombosis |
|  |  |  | Illness:I82.9 Embolism and thrombosis of unspecified vein |
|  |  |  | Illness:D69.2 Other nonthrombocytopenic purpura |
|  |  |  | Illness:I63.6 Cerebral infarction due to cerebral venous thrombosis- nonpyogenic |
|  |  |  | Illness:K64.5 Perianal venous thrombosis |
|  |  |  | Illness:I74.4 Embolism and thrombosis of arteries of extremities- unspecified |
|  |  |  | Illness:G08 Intracranial and intraspinal phlebitis and thrombophlebitis |
|  |  |  | Illness:O22.3 Deep phlebothrombosis in pregnancy |
|  |  |  | Illness:I82.3 Embolism and thrombosis of renal vein |
|  |  |  | Illness:O87.0 Superficial thrombophlebitis in the puerperium |
|  |  |  | Illness:Y44.4 Antithrombotic drugs [platelet-aggregation inhibitors] |
|  |  |  | Illness:O22.2 Superficial thrombophlebitis in pregnancy |
|  |  |  | Illness:Z92.1 Personal history of long-term (current) use of anticoagulants |
|  |  |  | Illness:T45.5 Anticoagulants |
|  |  |  | Illness:D65 Disseminated intravascular coagulation [defibrination syndrome] |
|  |  |  | Illness:D68.8 Other specified coagulation defects |
|  |  |  | Illness:D68.3 Haemorrhagic disorder due to circulating anticoagulants |
|  |  |  | Illness:Y44.2 Anticoagulants |
|  |  |  | Illness:D68.9 Coagulation defect- unspecified |
|  |  |  | Illness:D68.4 Acquired coagulation factor deficiency |
|  |  |  | Illness:O72.3 Postpartum coagulation defects |
|  |  |  | Illness:Y44.3 Anticoagulant antagonists- vitamin K and other coagulants |
|  |  |  | Illness:D68.2 Hereditary deficiency of other clotting factors |
|  |  |  | Illness:O88.2 Obstetric blood-clot embolism |
| Hepatorenal function | 6. Alanine aminotransferase (ALT)  7. Glucose | 24. Sodium | 30530: Sodium in urine |
|  |  | 25. Creatinine | 30510: Creatinine (enzymatic) in urine |
|  |  | 26. Body fat percentage | 23099: Body fat percentage |
|  |  | 27. Whole body fat mass | 23100: Whole body fat mass |
|  |  | 28. Whole body fat-free mass | 23101: Whole body fat-free mass |
|  |  | 29. Whole body water mass | 23102: Whole body water mass |
|  |  | 30. Body mass index | 23104: Body mass index (BMI) |
|  |  | 31. Basal metabolic rate | 23105: Basal metabolic rate |
|  |  | 32. Trunk fat percentage | 23127: Trunk fat percentage |
|  |  | 33. Trunk fat mass | 23128: Trunk fat mass |
|  |  | 34. Trunk fat-free mass | 23129: Trunk fat-free mass |
|  |  | 35. Trunk predicted mass | 23130: Trunk predicted mass |
|  |  | 36. Number of diabetes | 2443: Diabetes diagnosed by doctor |
|  |  |  | Illness:Z83.3 Family history of diabetes mellitus |
|  |  |  | Illness:N08.3 Glomerular disorders in diabetes mellitus |
|  |  |  | Illness:O24.0 Pre-existing diabetes mellitus- insulin-dependent |
|  |  |  | Illness:O24.4 Diabetes mellitus arising in pregnancy |
|  |  |  | Illness:Z13.1 Special screening examination for diabetes mellitus |
|  |  |  | Illness:E23.2 Diabetes insipidus |
|  |  |  | Illness:O24.9 Diabetes mellitus in pregnancy- unspecified |
|  |  |  | Illness:O24.1 Pre-existing diabetes mellitus- noninsulin-dependent |
|  |  |  | Illness:T38.3 Insulin and oral hypoglycaemic [antidiabetic] drugs |
|  |  |  | Illness:Y42.3 Insulin and oral hypoglycaemic [antidiabetic] drugs |
|  |  |  | Illness:E89.1 Postprocedural hypoinsulinaemia |
|  |  | 37. Number of kidney diseases | Illness:N18.3 Chronic kidney disease- stage 3 |
|  |  |  | Illness:N20.0 Calculus of kidney |
|  |  |  | Illness:N28.1 Cyst of kidney- acquired |
|  |  |  | Illness:N18.4 Chronic kidney disease- stage 4 |
|  |  |  | Illness:N18.2 Chronic kidney disease- stage 2 |
|  |  |  | Illness:N20.2 Calculus of kidney with calculus of ureter |
|  |  |  | Illness:C64 Malignant neoplasm of kidney- except renal pelvis |
|  |  |  | Illness:N28.8 Other specified disorders of kidney and ureter |
|  |  |  | Illness:R94.4 Abnormal results of kidney function studies |
|  |  |  | Illness:N28.9 Disorder of kidney and ureter- unspecified |
|  |  |  | Illness:N18.1 Chronic kidney disease- stage 1 |
|  |  |  | Illness:N18.5 Chronic kidney disease- stage 5 |
|  |  |  | Illness:Q61.9 Cystic kidney disease- unspecified |
|  |  |  | Illness:Q61.3 Polycystic kidney- unspecified |
|  |  |  | Illness:Z90.5 Acquired absence of kidney |
|  |  |  | Illness:S37.00 Injury of kidney (without open wound into cavity) |
|  |  |  | Illness:Q61.2 Polycystic kidney- adult type |
|  |  |  | Illness:N29.8 Other disorders of kidney and ureter in other diseases classified elsewhere |
|  |  |  | Illness:N27.9 Small kidney- unspecified |
|  |  |  | Illness:N26 Unspecified contracted kidney |
|  |  |  | Illness:N27.1 Small kidney- bilateral |
|  |  |  | Illness:Q63.0 Accessory kidney |
|  |  |  | Illness:N28.0 Ischaemia and infarction of kidney |
|  |  |  | Illness:S37.0 Injury of kidney |
|  |  |  | Illness:Q63.1 Lobulated- fused and horseshoe kidney |
|  |  |  | Illness:Z84.1 Family history of disorders of the kidney and ureter |
|  |  |  | Illness:C79.0 Secondary malignant neoplasm of kidney and renal pelvis |
|  |  |  | Illness:Q63.9 Congenital malformation of kidney- unspecified |
|  |  |  | Illness:Q63.2 Ectopic kidney |
|  |  |  | Illness:Q61.5 Medullary cystic kidney |
|  |  |  | Illness:N27.0 Small kidney- unilateral |
|  |  |  | Illness:Q63.8 Other specified congenital malformations of kidney |
|  |  |  | Illness:Y84.1 Kidney dialysis |
|  |  |  | Illness:Z94.0 Kidney transplant status |
|  |  |  | Illness:T86.1 Kidney transplant failure and rejection |
|  |  |  | Illness:N13.3 Other and unspecified hydronephrosis |
|  |  |  | Illness:N12 Tubulo-interstitial nephritis- not specified as acute or chronic |
|  |  |  | Illness:N13.2 Hydronephrosis with renal and ureteral calculous obstruction |
|  |  |  | Illness:N13.6 Pyonephrosis |
|  |  |  | Illness:N13.5 Kinking and stricture of ureter without hydronephrosis |
|  |  |  | Illness:N10 Acute tubulo-interstitial nephritis |
|  |  |  | Illness:N02.3 Diffuse mesangial proliferative glomerulonephritis |
|  |  |  | Illness:N13.1 Hydronephrosis with ureteral stricture- not elsewhere classified |
|  |  |  | Illness:N13.0 Hydronephrosis with ureteropelvic junction obstruction |
|  |  |  | Illness:N11.0 Nonobstructive reflux-associated chronic pyelonephritis |
|  |  |  | Illness:N05.3 Diffuse mesangial proliferative glomerulonephritis |
|  |  |  | Illness:N15.1 Renal and perinephric abscess |
|  |  |  | Illness:N11.9 Chronic tubulo-interstitial nephritis- unspecified |
|  |  |  | Illness:N11.1 Chronic obstructive pyelonephritis |
|  |  |  | Illness:N05.5 Diffuse mesangiocapillary glomerulonephritis |
|  |  |  | Illness:N04.5 Diffuse mesangiocapillary glomerulonephritis |
|  |  |  | Illness:N05.7 Diffuse crescentic glomerulonephritis |
|  |  |  | Illness:N04.2 Diffuse membranous glomerulonephritis |
|  |  |  | Illness:N02.2 Diffuse membranous glomerulonephritis |
|  |  |  | Illness:N05.2 Diffuse membranous glomerulonephritis |
|  |  |  | Illness:N11.8 Other chronic tubulo-interstitial nephritis |
|  |  |  | Illness:N14.1 Nephropathy induced by other drugs- medicaments and biological substances |
|  |  | 38. Number of liver diseases | Illness:K76.9 Liver disease- unspecified |
|  |  |  | Illness:C22.0 Liver cell carcinoma |
|  |  |  | Illness:Z94.4 Liver transplant status |
|  |  |  | Illness:O26.6 Liver disorders in pregnancy- childbirth and the puerperium |
|  |  |  | Illness:K70.3 Alcoholic cirrhosis of liver |
|  |  |  | Illness:K70.9 Alcoholic liver disease- unspecified |
|  |  |  | Illness:R94.5 Abnormal results of liver function studies |
|  |  |  | Illness:S36.10 Injury of liver or gallbladder (without open wound into cavity) |
|  |  |  | Illness:K76.0 Fatty (change of) liver- not elsewhere classified |
|  |  |  | Illness:K74.6 Other and unspecified cirrhosis of liver |
|  |  |  | Illness:K71.6 Toxic liver disease with hepatitis- not elsewhere classified |
|  |  |  | Illness:K76.8 Other specified diseases of liver |
|  |  |  | Illness:K75.0 Abscess of liver |
|  |  |  | Illness:K70.0 Alcoholic fatty liver |
|  |  |  | Illness:C78.7 Secondary malignant neoplasm of liver |
|  |  |  | Illness:R93.2 Abnormal findings on diagnostic imaging of liver and biliary tract |
|  |  |  | Illness:K75.9 Inflammatory liver disease- unspecified |
|  |  |  | Illness:K75.8 Other specified inflammatory liver diseases |
|  |  |  | Illness:K70.2 Alcoholic fibrosis and sclerosis of liver |
|  |  |  | Illness:S36.1 Injury of liver or gallbladder |
|  |  |  | Illness:Q44.6 Cystic disease of liver |
|  |  |  | Illness:K76.1 Chronic passive congestion of liver |
|  |  |  | Illness:K71.0 Toxic liver disease with cholestasis |
|  |  |  | Illness:K71.1 Toxic liver disease with hepatic necrosis |
|  |  |  | Illness:K71.5 Toxic liver disease with chronic active hepatitis |
|  |  |  | Illness:C22.4 Other sarcomas of liver |
|  |  |  | Illness:K71.8 Toxic liver disease with other disorders of liver |
| Blood pressure | 8. Mean arterial pressure | 39. Diastolic blood pressure | 4079: Diastolic blood pressure, automated reading |
|  |  | 40. Systolic blood pressure | 4080: Systolic blood pressure, automated reading |
|  |  | 41. Number of hypertension | Illness:I10 Essential (primary) hypertension |
|  |  |  | Illness:K76.6 Portal hypertension |
|  |  |  | Illness:O13 Gestational [pregnancy-induced] hypertension without significant proteinuria |
|  |  |  | Illness:O16 Unspecified maternal hypertension |
|  |  |  | Illness:Y52.5 Other antihypertensive drugs- not elsewhere classified |
|  |  |  | Illness:I27.2 Other secondary pulmonary hypertension |
|  |  |  | Illness:I27.0 Primary pulmonary hypertension |
|  |  |  | Illness:O10.0 Pre-existing essential hypertension complicating pregnancy- childbirth and the puerperium |
|  |  |  | Illness:I15.9 Seconday hypertension- unspecified |
|  |  |  | Illness:T46.5 Other antihypertensive drugs- not elsewhere classified |
|  |  |  | Illness:G93.2 Benign intracranial hypertension |
|  |  |  | Illness:I15.0 Renovascular hypertension |
|  |  |  | Illness:O11 Pre-existing hypertensive disorder with superimposed proteinuria |
| Inflammatory | 9. While blood cells  10. Lymphocytes  11. Interleukin-6  12. C-reactive protein  13. Procalcitonin | 42. White blood cell count | 30000: White blood cell (leukocyte) count |
|  |  | 43. Lymphocyte count | 30120: Lymphocyte count |
|  |  | 44. Neutrophill count | 30140: Neutrophill count |
|  |  | 45. Eosinophill count | 30150: Eosinophill count |
|  |  | 46. Basophill count | 30160: Basophill count |
|  |  | 47. Lymphocyte percentage | 30180: Lymphocyte percentage |
|  |  | 48. Monocyte percentage | 30190: Monocyte percentage |
|  |  | 49. Neutrophill percentage | 30200: Neutrophill percentage |
|  |  | 50. Eosinophill percentage | 30210: Eosinophill percentage |
|  |  | 51. Basophill percentage | 30220: Basophill percentage |
|  |  | 52. Number of inflammatory | Illness:T82.7 Infection and inflammatory reaction due to other cardiac and vascular devices- implants and grafts |
|  |  |  | Illness:T84.5 Infection and inflammatory reaction due to internal joint prosthesis |
|  |  |  | Illness:T84.7 Infection and inflammatory reaction due to other internal orthopaedic prosthetic devices- implants and grafts |
|  |  |  | Illness:I83.1 Varicose veins of lower extremities with inflammation |
|  |  |  | Illness:N73.9 Female pelvic inflammatory disease- unspecified |
|  |  |  | Illness:T85.7 Infection and inflammatory reaction due to other internal prosthetic devices- implants and grafts |
|  |  |  | Illness:H30.9 Chorioretinal inflammation- unspecified |
|  |  |  | Illness:T83.5 Infection and inflammatory reaction due to prosthetic device- implant and graft in urinary system |
|  |  |  | Illness:T84.6 Infection and inflammatory reaction due to internal fixation device [any site] |
|  |  |  | Illness:T39.3 Other nonsteroidal anti-inflammatory drugs [NSAID] |
|  |  |  | Illness:Y45.3 Other nonsteroidal anti-inflammatory drugs [NSAID] |
|  |  |  | Illness:K75.8 Other specified inflammatory liver diseases |
|  |  |  | Illness:G61.8 Other inflammatory polyneuropathies |
|  |  |  | Illness:N76.8 Other specified inflammation of vagina and vulva |
|  |  |  | Illness:G09 Sequelae of inflammatory diseases of central nervous system |
|  |  |  | Illness:I83.2 Varicose veins of lower extremities with both ulcer and inflammation |
|  |  |  | Illness:H00.0 Hordeolum and other deep inflammation of eyelid |
|  |  |  | Illness:T82.6 Infection and inflammatory reaction due to cardiac valve prosthesis |
|  |  |  | Illness:H04.4 Chronic inflammation of lachrymal passages |
|  |  |  | Illness:H05.0 Acute inflammation of orbit |
|  |  |  | Illness:N73.8 Other specified female pelvic inflammatory diseases |
|  |  |  | Illness:N48.2 Other inflammatory disorders of penis |
|  |  |  | Illness:E79.0 Hyperuricaemia without signs of inflammatory arthritis and tophaceous disease |
|  |  |  | Illness:M46.82 Other specified inflammatory spondylopathies (Cervical region) |
|  |  |  | Illness:H40.4 Glaucoma secondary to eye inflammation |
|  |  |  | Illness:H04.3 Acute and unspecified inflammation of lachrymal passages |
|  |  |  | Illness:L81.0 Postinflammatory hyperpigmentation |
|  |  |  | Illness:Y56.0 Local antifungal- anti-infective and anti-inflammatory drugs- not elsewhere classified |
|  |  |  | Illness:M46.80 Other specified inflammatory spondylopathies (Multiple sites in spine) |
|  |  |  | Illness:N71.1 Chronic inflammatory disease of uterus |
|  |  |  | Illness:N74.8 Female pelvic inflammatory disorders in other diseases classified elsewhere |
|  |  |  | Illness:H01.8 Other specified inflammation of eyelid |
|  |  |  | Illness:T83.6 Infection and inflammatory reaction due to prosthetic device- implant and graft in genital tract |
|  |  |  | Illness:H32.0 Chorioretinal inflammation in infectious and parasitic diseases classified elsewhere |
|  |  |  | Illness:J68.2 Upper respiratory inflammation due to chemicals- gases- fumes and vapours- not elsewhere classified |
|  |  |  | Illness:N41.9 Inflammatory disease of prostate- unspecified |
|  |  |  | Illness:M46.9 Inflammatory spondylopathy- unspecified |
|  |  |  | Illness:M46.92 Inflammatory spondylopathy- unspecified (Cervical region) |
|  |  |  | Illness:N49.2 Inflammatory disorders of scrotum |
|  |  |  | Illness:M46.96 Inflammatory spondylopathy- unspecified (Lumbar region) |
|  |  |  | Illness:N72 Inflammatory disease of cervix uteri |
|  |  |  | Illness:K75.9 Inflammatory liver disease- unspecified |
|  |  |  | Illness:R65.1 Systemic Inflammatory Response Syndrome of infectious origin with organ failure |
|  |  |  | Illness:N61 Inflammatory disorders of breast |
|  |  |  | Illness:R65.2 Systemic Inflammatory Response Syndrome of non-infectious origin without organ failure |
|  |  |  | Illness:N71.9 Inflammatory disease of uterus- unspecified |
|  |  |  | Illness:M46.99 Inflammatory spondylopathy- unspecified (Site unspecified) |
|  |  |  | Illness:G61.9 Inflammatory polyneuropathy- unspecified |
|  |  |  | Illness:R65.0 Systemic Inflammatory Response Syndrome of infectious origin without organ failure |
|  |  |  | Illness:M06.40 Inflammatory polyarthropathy (Multiple sites) |
|  |  |  | Illness:M06.49 Inflammatory polyarthropathy (Site unspecified) |
|  |  |  | Illness:M46.97 Inflammatory spondylopathy- unspecified (Lumbosacral region) |
|  |  |  | Illness:K10.2 Inflammatory conditions of jaws |
|  |  |  | Illness:R65.9 Systemic Inflammatory Response Syndrome- unspecified |
|  |  |  | Illness:H01.9 Inflammation of eyelid- unspecified |
|  |  |  | Illness:M46.90 Inflammatory spondylopathy- unspecified (Multiple sites in spine) |
|  |  |  | Illness:M46.98 Inflammatory spondylopathy- unspecified (Sacral and sacrococcygeal region) |
|  |  |  | Illness:M06.4 Inflammatory polyarthropathy |
|  |  |  | Illness:M46.94 Inflammatory spondylopathy- unspecified (Thoracic region) |
|  |  |  | Illness:G72.4 Inflammatory myopathy- not elsewhere classified |
|  |  |  | Illness:N49.1 Inflammatory disorders of spermatic cord- tunica vaginalis and vas deferens |
|  |  |  | Illness:M06.45 Inflammatory polyarthropathy (Pelvic region and thigh) |
|  |  | 53. Number of calcium and phosphorus metabolic diseases | Illness:E83.5 Disorders of calcium metabolism |
|  |  |  | Illness:E07.0 Hypersecretion of calcitonin |
|  |  |  | Illness:E83.3 Disorders of phosphorus metabolism |
| Cardiovascular function | 14. Troponin | 54. Pulse rate | 102: Pulse rate, automated reading |
|  |  | 55. Number of heart diseases | Illness:Z13.6 Special screening examination for cardiovascular disorders |
|  |  |  | Illness:R94.3 Abnormal results of cardiovascular function studies |
|  |  |  | Illness:I42.0 Dilated cardiomyopathy |
|  |  |  | Illness:I42.2 Other hypertrophic cardiomyopathy |
|  |  |  | Illness:I42.1 Obstructive hypertrophic cardiomyopathy |
|  |  |  | Illness:I42.8 Other cardiomyopathies |
|  |  |  | Illness:Z03.5 Observation for other suspected cardiovascular diseases |
|  |  |  | Illness:I25.0 Atherosclerotic cardiovascular disease- so described |
|  |  |  | Illness:I25.5 Ischaemic cardiomyopathy |
|  |  |  | Illness:I42.6 Alcoholic cardiomyopathy |
|  |  |  | Illness:I42.5 Other restrictive cardiomyopathy |
|  |  |  | Illness:I51.7 Cardiomegaly |
|  |  |  | Illness:I42.9 Cardiomyopathy- unspecified |
|  |  |  | Illness:I51.6 Cardiovascular disease- unspecified |
|  |  |  | Illness:I43.1 Cardiomyopathy in metabolic diseases |
|  |  |  | Illness:R57.0 Cardiogenic shock |
|  |  |  | 6150: Vascular/heart problems diagnosed by doctor |
|  |  |  | Illness:Z82.4 Family history of ischaemic heart disease and other diseases of the circulatory system |
|  |  |  | Illness:I25.9 Chronic ischaemic heart disease- unspecified |
|  |  |  | Illness:I25.1 Atherosclerotic heart disease |
|  |  |  | Illness:I24.9 Acute ischaemic heart disease- unspecified |
|  |  |  | Illness:I25.8 Other forms of chronic ischaemic heart disease |
|  |  |  | Illness:Z95.2 Presence of prosthetic heart valve |
|  |  |  | Illness:Z95.3 Presence of xenogenic heart valve |
|  |  |  | Illness:O68.0 Labour and delivery complicated by foetal heart rate anomaly |
|  |  |  | Illness:Z95.4 Presence of other heart-valve replacement |
|  |  |  | Illness:R93.1 Abnormal findings on diagnostic imaging of heart and coronary circulation |
|  |  |  | Illness:I50.0 Congestive heart failure |
|  |  |  | Illness:I51.8 Other ill-defined heart diseases |
|  |  |  | Illness:I27.9 Pulmonary heart disease- unspecified |
|  |  |  | Illness:Q24.8 Other specified congenital malformations of heart |
|  |  |  | Illness:O68.2 Labour and delivery complicated by foetal heart rate anomaly with meconium in amniotic fluid |
|  |  |  | Illness:I25.3 Aneurysm of heart |
|  |  |  | Illness:R00.8 Other and unspecified abnormalities of heart beat |
|  |  |  | Illness:I24.8 Other forms of acute ischaemic heart disease |
|  |  |  | Illness:T82.0 Mechanical complication of heart valve prosthesis |
|  |  |  | Illness:I45.5 Other specified heart block |
|  |  |  | Illness:I11.9 Hypertensive heart disease without (congestive) heart failure |
|  |  |  | Illness:I11.0 Hypertensive heart disease with (congestive) heart failure |
|  |  |  | Illness:S26.80 Other injuries of heart (without open wound into thoracic cavity) |
|  |  |  | Illness:I52.8 Other heart disorders in other diseases classified elsewhere |
|  |  |  | Illness:Q24.6 Congenital heart block |
|  |  |  | Illness:I09.9 Rheumatic heart disease- unspecified |
|  |  |  | Illness:I13.9 Hypertensive heart and renal disease- unspecified |
|  |  |  | Illness:I00 Rheumatic fever without mention of heart involvement |
|  |  |  | Illness:Q24.9 Congenital malformation of the heart- unspecified |
|  |  |  | Illness:I13.2 Hypertensive heart and renal disease with both (congestive) heart failure and renal failure |
|  |  |  | Illness:I27.8 Other specified pulmonary heart diseases |
|  |  |  | Illness:I50.9 Heart failure- unspecified |
|  |  |  | Illness:I51.9 Heart disease- unspecified |
|  |  |  | Illness:Z94.1 Heart transplant status |
|  |  |  | Illness:D15.1 Heart |
|  |  |  | Illness:T67.1 Heat syncope |
|  |  |  | Illness:R55 Syncope and collapse |
|  |  |  | Illness:I49.9 Cardiac arrhythmia- unspecified |
|  |  |  | Illness:I47.0 Reentry ventricular arrhythmia |
|  |  |  | Illness:I49.8 Other specified cardiac arrhythmias |
|  |  |  | Illness:R00.2 Palpitations |
|  |  |  | 2316: Wheeze or whistling in the chest in last year |
|  |  |  | 3090: Used an inhaler for chest within last hour |
|  |  |  | Illness:R07.3 Other chest pain |
|  |  |  | Illness:S22.50 Flail chest (closed) |
|  |  |  | Illness:M95.4 Acquired deformity of chest and rib |
|  |  |  | 2335: Chest pain or discomfort |
|  |  |  | 3606: Chest pain or discomfort walking normally |
|  |  |  | 3616: Chest pain due to walking ceases when standing still |
|  |  |  | 3751: Chest pain or discomfort when walking uphill or hurrying |
|  |  |  | Illness:R07.4 Chest pain- unspecified |
|  |  |  | Illness:R07.1 Chest pain on breathing |

**Supplementary Table 4. Features selected for Cohort 1**

| **Fetures** | **XGBoost** | **GBDT** | **RF** | **LR** | **SVM** | **Selected** |
| --- | --- | --- | --- | --- | --- | --- |
| Age | Yes | Yes | Yes | Yes | Yes | Yes |
| OsSats | Yes | Yes | Yes | Yes | Yes |  |
| MAP | Yes | Yes | Yes | Yes | Yes |  |
| Ddimer | Yes | Yes | Yes | Yes | Yes |  |
| Glucose | Yes | Yes | Yes | Yes | Yes |  |
| WBC | Yes | Yes | Yes | Yes | Yes |  |
| Lympho | Yes | Yes | Yes | Yes | Yes |  |
| IL6 | Yes | Yes | Yes | Yes | Yes |  |
| CrctProtein | Yes | Yes | Yes | Yes | Yes |  |
| Procalcitonin | Yes | Yes | Yes | Yes | Yes |  |
| Troponin | Yes | Yes | Yes | No | Yes |  |
| Plts | No | No | Yes | Yes | Yes |  |
| INR | Yes | Yes | Yes | No | No |  |
| ALT | No | Yes | No | Yes | Yes |  |
| Temp | Yes | No | Yes | No | No | No |
| BUN | No | No | No | Yes | Yes |  |
| Creatine | No | No | No | Yes | Yes |  |
| Sodium | Yes | No | No | No | No |  |
| AST | No | No | No | Yes | No |  |
| Ferritin | No | No | No | Yes | No |  |

Note: “Yes”(”No”) means that the feature is (not) selected by base model.

**Supplementary Table 5. The performance comparison of each predictive model after adopting different imputation methods on Cohort 1**

| **GBDT** | **KNN** | **Mean** | **Median** | **Chained equation[2]** | **Constant (0)** |
| --- | --- | --- | --- | --- | --- |
| Accuracy (SD) | 0.864(0.005) | 0.840(0.005) | 0.838(0.005) | 0.849(0.005) | 0.842(0.005) |
| AUC (SD) | **0.900(0.005)** | 0.854(0.007) | 0.855(0.007) | 0.875(0.006) | 0.857(0.007) |
| Precisions (SD) | 0.774(0.018) | 0.745(0.019) | 0.741(0.020) | 0.748(0.018) | 0.753(0.019) |
| Recall (SD) | 0.625(0.016) | 0.520(0.019) | 0.519(0.019) | 0.572(0.019) | 0.525(0.019) |
| **XGBoost** | **KNN** | **Mean** | **Median** | **Chained equation** | **Constant (0)** |
| Accuracy (SD) | 0.864(0.005) | 0.835(0.005) | 0.835(0.005) | 0.845(0.005) | 0.836(0.004) |
| AUC (SD) | **0.904(0.005)** | 0.853(0.007) | 0.852(0.006) | 0.874(0.006) | 0.853(0.006) |
| Precisions (SD) | 0.805(0.019) | 0.759(0.020) | 0.759(0.018) | 0.763(0.018) | 0.764(0.018) |
| Recall (SD) | 0.581(0.017) | 0.474(0.019) | 0.475(0.019) | 0.530(0.019) | 0.475(0.019) |
| **RF** | **KNN** | **Mean** | **Median** | **Chained equation** | **Constant (0)** |
| Accuracy (SD) | 0.861(0.005) | 0.839(0.004) | 0.837(0.004) | 0.849(0.005) | 0.838(0.005) |
| AUC (SD) | **0.900(0.005)** | 0.855(0.007) | 0.853(0.006) | 0.877(0.007) | 0.856(0.006) |
| Precisions (SD) | 0.791(0.019) | 0.762(0.019) | 0.760(0.019) | 0.769(0.018) | 0.765(0.019) |
| Recall (SD) | 0.588(0.018) | 0.493(0.020) | 0.486(0.020) | 0.544(0.021) | 0.484(0.020) |
| **LR** | **KNN** | **Mean** | **Median** | **Chained equation** | **Constant (0)** |
| Accuracy (SD) | 0.847(0.004) | 0.829(0.005) | 0.827(0.005) | 0.836(0.004) | 0.824(0.005) |
| AUC (SD) | **0.870(0.006)** | 0.828(0.007) | 0.824(0.007) | 0.844(0.007) | 0.820(0.007) |
| Precisions (SD) | 0.764(0.018) | 0.740(0.018) | 0.738(0.019) | 0.749(0.017) | 0.732(0.019) |
| Recall (SD) | 0.542(0.019) | 0.458(0.018) | 0.452(0.017) | 0.494(0.019) | 0.436(0.018) |
| **SVM** | **KNN** | **Mean** | **Median** | **Chained equation** | **Constant (0)** |
| Accuracy (SD) | 0.855(0.006) | 0.827(0.005) | 0.825(0.005) | 0.836(0.005) | 0.827(0.005) |
| AUC (SD) | **0.890(0.005)** | 0.843(0.007) | 0.838(0.007) | 0.864(0.006) | 0.840(0.007) |
| Precisions (SD) | 0.738(0.020) | 0.703(0.019) | 0.696(0.019) | 0.713(0.019) | 0.701(0.019) |
| Recall (SD) | 0.628(0.021) | 0.501(0.018) | 0.498(0.018) | 0.550(0.018) | 0.508(0.019) |
| **EM** | **KNN** | **Mean** | **Median** | **Chained equation** | **Constant (0)** |
| Accuracy (SD) | 0.867(0.005) | 0.841(0.005) | 0.840(0.004) | 0.850(0.005) | 0.842(0.005) |
| AUC (SD) | **0.908(0.005)** | 0.862(0.007) | 0.861(0.006) | 0.881(0.006) | 0.862(0.006) |
| Precisions (SD) | 0.806(0.019) | 0.773(0.019) | 0.772(0.018) | 0.771(0.018) | 0.779(0.019) |
| Recall (SD) | 0.600(0.016) | 0.493(0.018) | 0.490(0.019) | 0.549(0.018) | 0.492(0.019) |

Note: We use the mean, median, chained equation[2] and constant (0) to impute the missing data in Cohort 1 respectively, and evaluate each model with the data processed by different imputation methods.

**Supplementary Table 6. Features selected for Cohort 2**

| **Features** | **XGBoost** | **GBDT** | **RF** | **LR** | **SVM** | **Selected** |
| --- | --- | --- | --- | --- | --- | --- |
| Age | Yes | Yes | Yes | Yes | Yes | Yes |
| Number of lung diseases | Yes | Yes | Yes | Yes | Yes |  |
| Number of hypertension | Yes | Yes | Yes | Yes | Yes |  |
| Monocyte percentage | Yes | Yes | Yes | Yes | Yes |  |
| Nucleated red blood cell count | Yes | Yes | Yes | No | Yes |  |
| Number of thrombotic diseases | Yes | Yes | Yes | Yes | No |  |
| Body fat percentage | Yes | Yes | No | Yes | Yes |  |
| Whole body fat mass | Yes | Yes | Yes | No | Yes |  |
| Body mass index | Yes | No | Yes | Yes | Yes |  |
| Number of liver diseases | Yes | Yes | Yes | Yes | No |  |
| Red blood cell distribution width | No | No | Yes | Yes | Yes |  |
| Platelet crit | Yes | Yes | No | Yes | No |  |
| Creatinine | Yes | Yes | Yes | No | No |  |
| Whole body water mass | Yes | No | No | Yes | Yes |  |
| Trunk fat mass | Yes | No | Yes | Yes | No |  |
| Number of diabetes | Yes | Yes | No | Yes | No |  |
| Pulse rate | Yes | Yes | No | Yes | No |  |
| Red blood cell count | No | Yes | Yes | No | No | No |
| Nucleated red blood cell percentage | Yes | No | No | No | Yes |  |
| Whole body fat-free mass | No | No | Yes | No | Yes |  |
| Basal metabolic rate | No | Yes | No | No | Yes |  |
| Trunk fat-free mass | No | Yes | No | No | Yes |  |
| Trunk predicted mass | No | Yes | No | No | Yes |  |
| White blood cell count | Yes | Yes | No | No | No |  |
| Lymphocyte count | No | No | Yes | No | Yes |  |
| Neutrophill count | No | No | Yes | Yes | No |  |
| Lymphocyte percentage | No | No | No | Yes | Yes |  |
| Basophill percentage | Yes | Yes | No | No | No |  |
| Number of heart diseases | Yes | No | Yes | No | No |  |
| Haematocrit percentage | No | No | Yes | No | No |  |
| Mean corpuscular haemoglobin | No | No | Yes | No | No |  |
| Reticulocyte percentage | No | No | No | No | Yes |  |
| Reticulocyte count | No | No | No | No | Yes |  |
| Mean sphered cell volume | No | No | Yes | No | No |  |
| Immature reticulocyte fraction | No | Yes | No | No | No |  |
| High light scatter reticulocyte percentage | No | No | No | No | Yes |  |
| High light scatter reticulocyte count | No | Yes | No | No | No |  |
| Platelet count | No | No | No | Yes | No |  |
| Mean platelet volume | No | No | No | No | Yes |  |
| Sodium | No | Yes | No | No | No |  |
| Number of kidney diseases | No | No | No | Yes | No |  |
| Diastolic blood pressure | Yes | No | No | No | No |  |
| Eosinophill count | No | Yes | No | No | No |  |
| Basophill count | No | Yes | No | No | No |  |
| Neutrophill percentage | No | No | No | No | Yes |  |
| Number of calcium and phosphorus metabolic diseases | No | No | Yes | No | No |  |
| Haemoglobin concentration | No | No | No | No | No |  |
| Mean corpuscular volume | No | No | No | No | No |  |
| Mean corpuscular haemoglobin concentration | No | No | No | No | No |  |
| Mean reticulocyte volume | No | No | No | No | No |  |
| Platelet distribution width | No | No | No | No | No |  |
| Trunk fat percentage | No | No | No | No | No |  |
| Systolic blood pressure | No | No | No | No | No |  |
| Eosinophill percentage | No | No | No | No | No |  |
| Number of inflammatory | No | No | No | No | No |  |

Note: “Yes”(”No”) means that the feature is (not) selected by base model.

**Supplementary Table 7. Comparison of several predictive models**

| **Authors** | **Elza Rechtman et al.[3]** | **David J. Altschul et al.[1]** | **Noam Barda et al.[4]** | **Li Yan et al.[5]** | **Wenhua Liang et al.[6]** | **Wanshan Ning et al.[7]** | **Shuo Wang et al.[8]** |
| --- | --- | --- | --- | --- | --- | --- | --- |
| Selected features | Age, gender, race, ethnicity, smoking status, BMI, heart rate, temperature, respiratory rate, oxygen saturation, chronic kidney disease, asthma, COPD, hypertension, diabetes, HIV, cancer | Age, Oxygen saturation, MAP, BUN, C-reactive protein, INR | Age, Sex, Pack years, COPD, Number of wheezing / dyspnea diagnoses, Albumin, Red cell distribution width, C-Reactive Peptide, Urea, Lymphocyte, Chloride, Creatinine, High Density Lipoprotein, Duration of hospitalizations, Count of hospitalizations, Count of ambulance rides, Count of Sulfonamide dispenses, Count of Anticholinergic dispenses, Count of Glucocorticoid dispenses, Chronic Respiratory Disease, Cardiovascular Disease, Diabetes, Malignancy, Hypertension | Lactic dehydrogenase, lymphocytes, high-sensitivity C-reactive protein | X-ray abnormalities, age, dyspnea, COPD, number of comorbidities, cancer history, neutrophil/lymphocytes ratio, lactate dehydrogenase, direct bilirubin, creatine kinase | 1. CT images  2. clinical features (CFs) data : 130 types from 9 categories, including basic information, routine blood test, inflammation test, blood coagulation test, biochemical test, immune cell typing, cytokine profile test, autoimmune test and routine urine test | 1. CT images (diagnosis);  2. Age, DL feature-22, DL feature-44 (prognosis) |
| Feature selection method | Feature selection is not performed | 1. Multiple logistic regression  2. Backward stepwise bootstrap regression model | 1. Cumulative information gain  2. Clinicians manually select features  3. Features that have been reported to be related to severe COVID-19 | Feature importance | LASSO algorithm | Feature selection is not performed | Stepwise prognostic feature selection |
| Number of patients | 8770 | 4711 | 1,050,000 non-COVID-19 patients (Baseline model establishment)  4179 COVID-19 patients | 485 | Training cohort : 1590  Validation set 1 : 940  Validation set 2 : 380  Validation set 3 : 73 | Cohort 1 : 1170 (649 patients with COVID-19)  Cohort 2 : 351 (245 patients with COVID-19) | 4106 (CT-EGGR dataset),  1266 (COVID-19 dataset) |
| Predictive model | Xgboost | COVID-19 severity score | LightGBM | XGBoost | Deep Learning Survival Cox model | Hybrid learning for unbiased prediction of COVID-19 patients (HUST-19) | 1. Deep learning model (diagnosis);  2. Multivariate Cox proportional hazard model (prognosis) |
| Clinical outcomes | Death or alive | Death or alive | Death or alive | Death or alive | Critical illness or not | 1. Mild or regular form (type I) and severe or critically ill form (type II)) (morbidity);  2. Death or alive (mortality) | 1. COVID-19 or other pneumonia (diagnosis)  2. Hospital stay time (prognosis) |
| Model performance evaluation | AUC : 0.86 | AUC : 0.824 (In the derivation cohort);  AUC : 0.798 (In the validation cohort) | Overall AUC : 0.943;  Threshold 10%:  Recall : 0.71  Precision : 0.30;  Threshold 5%:  Recall : 0.88  Precision : 0.20 | AUC : 0.9506 | AUC : 0.911 (In the internal validation cohort)  AUC : 0.881 (In the external validation set 1)  AUC : 0.819 (In the external validation set 2)  AUC : 0.967 (In the external validation set 3) | 1. Morbidity prediction (AUC): 0.978 (controls in cohort 1), 0.921 (type  I in cohort 1), 0.931 (type II in cohort 1) 0.944 (controls in cohort 2), 0.860 (type  I in cohort 2), 0.884 (type II in cohort 1)  2. Mortality prediction (AUC): 0.856 (total) | 1. Diagnosis (AUC): 0.90 (In the training set), 0.87 (In the validation set 1), 0.88 (In the validation set 2. Prognosis : (log-rank test): p<0.0001 (In the training set), p=0.013 (In the validation sets 3), p=0.014 (In the validation sets 4) |

**Supplementary Table 8. Performance comparison of predictive models**

| **Model** | **Number of patients (Death / Total)** | **AUC** | **Precision (when recall reaches 0.71)** | **Precision (when recall reaches 0.88)** |
| --- | --- | --- | --- | --- |
| EM | 1148 / 4711 | 0.908 | 0.74 | 0.57 |
| LightGBM | 143 /4179 | 0.943 | 0.30 | 0.20 |

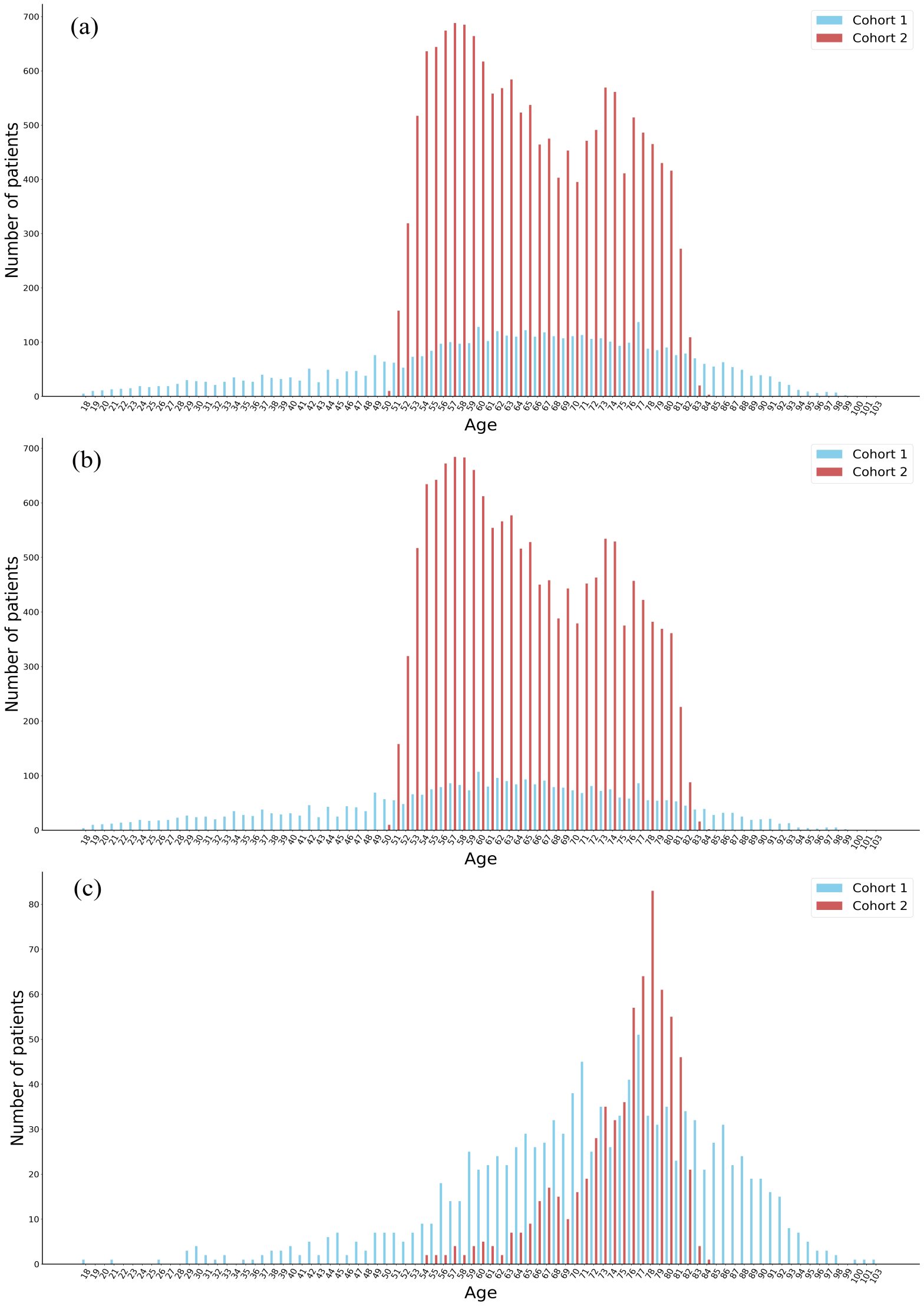

Supplementary Figure 1. Age distribution of patients in Cohor 1 and Cohort 2. (a) All patients. (b) Alive patients. (c) Dead patients.

**Reference**

1.	Altschul DJ, Unda SR, Benton J, de la Garza Ramos R, Cezayirli P, Mehler M, et al. A novel severity score to predict inpatient mortality in COVID-19 patients. Scientific Reports. 2020;10(1):16726.

2.	White IR, Royston P, Wood AM. Multiple imputation using chained equations: Issues and guidance for practice. Statistics in Medicine. 2011;30(4):377-99.

3.	Rechtman E, Curtin P, Navarro E, Nirenberg S, Horton MK. Vital signs assessed in initial clinical encounters predict COVID-19 mortality in an NYC hospital system. Scientific Reports. 2020;10(1):21545.

4.	Barda N, Riesel D, Akriv A, Levy J, Finkel U, Yona G, et al. Developing a COVID-19 mortality risk prediction model when individual-level data are not available. Nature Communications. 2020;11(1):4439.

5.	Yan L, Zhang H-T, Goncalves J, Xiao Y, Wang M, Guo Y, et al. An interpretable mortality prediction model for COVID-19 patients. Nature Machine Intelligence. 2020;2(5):283-8.

6.	Liang W, Yao J, Chen A, Lv Q, Zanin M, Liu J, et al. Early triage of critically ill COVID-19 patients using deep learning. Nature Communications. 2020;11(1):3543.

7.	Ning W, Lei S, Yang J, Cao Y, Jiang P, Yang Q, et al. Open resource of clinical data from patients with pneumonia for the prediction of COVID-19 outcomes via deep learning. Nature Biomedical Engineering. 2020;4(12):1197-207.

8.	Wang S, Zha Y, Li W, Wu Q, Li X, Niu M, et al. A fully automatic deep learning system for COVID-19 diagnostic and prognostic analysis. European Respiratory Journal. 2020;56(2):2000775.
